# Avidity of anti-pertussis toxin antibodies is associated with symptomatic *Bordetella pertussis* infection in a novel controlled human infection model

**DOI:** 10.64898/2026.06.17.26355173

**Authors:** Carlos Espinosa-Vinals, Hala Obeid, Kara L Redden, May ElSherif, Kathryn M. Edwards, Scott Halperin, Bahaa Abu-Raya

## Abstract

**Background:** The association between functional antibody responses following *Bordetella pertussis* infection and symptomatic disease remains unclear. We characterized the maturation of anti-pertussis toxin (PT) IgG avidity after human challenge with *B. pertussis* and determined its association with symptomatic infection.

**Methods:** Healthy adults were intranasally inoculated with live *B. pertussis* organisms in a controlled human infection model and monitored for development of pertussis symptoms (NCT05136599). Serum samples were collected one day before inoculation and at 14, 28, 56, 180, and 365 days post-challenge. Anti-PT IgG avidity was tested using a titration of ammonium isothiocyanate (the bond-breaking agent) to quantify a wide range of antibody avidities from low to very-high. Associations between covariates and avidity were examined using linear regression models, and high-dimensional analyses were used to integrate all data.

**Findings:** Anti-PT IgG avidity increased in both symptomatic (n=20) and asymptomatic (n=10) participants after the challenge, reached maximum levels at day 56, and then declined through day 365. Symptomatic participants developed significantly higher levels of high- and very high-avidity anti-PT antibodies at 28, 56, 180, and 365 days post-challenge compared with those who remained asymptomatic. In multivariate analyses, symptomatic infection was associated with higher levels of high and very high avidity anti-PT IgG at day□180 and□365 after challenge. Distinct avidity profiles in symptomatic vs asymptomatic participants emerged at day□28 onwards, with the former group having higher levels of antibodies with higher avidities. However, levels of medium-high, high and very high avidity antibodies in symptomatic participants were lower at day 365 after challenge compared to their peak levels.

**Interpretation:** Anti-PT IgG avidity was associated with symptomatic *B. pertussis* infection and thus may serve as a surrogate of clinical disease outcome. These results highlight that antibody avidity provides an additional functional assay besides antibody quantitation to dissect immune responses to pertussis. Further investigation of anti-PT IgG avidity should be pursued in natural pertussis outbreaks to determine whether it might be used to differentiate symptomatic from asymptomatic infections for epidemiologic purposes.

**Funding:** Canadian Association for Immunization Research Evaluation and Education.

**Research in context:** *Evidence before this study:* The relationship between antibody immune responses to *B. pertussis* infection and clinical disease is incompletely characterized. While antibody quantification is commonly measured, functional assessments of antibody can provide additional information. Antibody avidity, reflecting the binding strength of antibodies to their antigen, may provide additional insight into antibody function. While *B. pertussis* expresses several antigens, pertussis toxin (PT) is specific to *B. pertussis* and is a pivotal virulence factor. For this reason, it has been chosen as the target for high-resolution avidity profiling in our study characterizing antibody immune responses after controlled human challenge with *B. pertussis*. Traditionally, avidity has been measured based on a comparison of antibody levels with and without the addition of a single concentration of ammonium isothiocyanate (as a bond-breaking agent), leading to an arbitrary separation into “low” and “high” avidity antibodies. However, relying on this traditional approach of using a single concentration of ammonium isothiocyanate provides more limited insight into avidity quantification and maturation. In addition, studies assessing avidity after *B. pertussis* infection have been limited, cross-sectional, and lacked well-defined characterization of *B. pertussis* exposure and clinical disease outcomes. Consequently, the potential relevance of anti-PT IgG avidity as a surrogate of pertussis clinical disease outcome remains insufficiently defined.

*Added value of this study:* We used sera from participants enrolled in a controlled human infection model with a well-defined exposure to a standardized *B. pertussis* challenge dose, frequent longitudinal sampling, and clearly defined clinical disease outcomes (symptomatic *versus* asymptomatic infection). We then applied a novel analytical approach that measures anti-PT IgG avidity across a wide spectrum of binding strengths. We clearly demonstrated higher anti-PT IgG avidity in symptomatic participants, compared with those who remained asymptomatic after inoculation. Anti-PT IgG avidity peaked at day 56 post-challenge, and then avidity antibodies subsequently declined through day 365 in symptomatic participants. These findings suggest that high-resolution avidity profiling provides additional detail about the immune response after asymptomatic and asymptomatic infections.

*Implications of all the available evidence:* Anti-PT IgG avidity emerges from our findings as a clinically relevant, functional component of the antibody response to *B. pertussis* infection. Avidity may contribute useful information for differentiating clinical disease outcomes and interpreting infection history. Additional studies are needed to confirm these findings after natural pertussis outbreaks and exposures.

## Introduction

The association between antibody immune response and symptomatic pertussis disease remains unclear. Efforts to identify an immune correlate of protection (CoP) —a specific function or level of immune system component(s) or markers that are correlated with protection against pertussis— have been limited and produced inconsistent results (1–3). Collectively, these findings indicate that anti-*B. pertussis* IgG levels (*i.e.* quantity) may not fully account for protection from pertussis and that evaluating the function (*i.e.* quality) of these antibodies may provide additional insight into the association with clinical outcomes after exposure to *B. pertussis*. One functional characteristic of antibodies is avidity, which measures the strength of antibody binding to its antigen. However, studies assessing the relationship between antibody avidity following natural *B. pertussis* infection and clinical disease outcomes are scarce (4).

Controlled human infection models (CHIMs) provide an extraordinary opportunity to examine immune responses in the context of precisely defined exposure and prospectively assessed clinical outcomes. Our group has recently established a unique CHIM with clearly defined clinical disease endpoints of both symptomatic *vs* asymptomatic infection.

In this study, we employed a novel approach to measure avidity maturation of anti-PT IgG following challenge and compared the responses in both symptomatic and asymptomatic infection. We focused on PT-specific antibody responses because PT is a critical virulence factor involved in immune modulation and *B. pertussis* pathogenesis, is uniquely specific to *B. pertussis*, and is included in all licensed acellular pertussis (aP) vaccines (5), making it a relevant and widely used antigen for immunological assessment.

## Materials and Methods

### Study cohort

This study was conducted in participants in a *B. pertussis* CHIM at the Canadian Center for Vaccinology (Halifax, Nova Scotia, Canada), as reported by the accompanying article by ElSherif and colleagues (6). Only participants with anti-PT IgG levels below 20 EU/mL at screening visit were eligible for enrollment. Participants were challenged with doses ranging from 5×10□ CFU to 5×10□ CFU of *B. pertussis.* Subjects were monitored for solicited systemic symptoms (systemic: fever, fatigue, malaise; respiratory: nasal congestion, rhinorrhea, sneezing, sore throat, watery eyes, red eyes, cough) and classified into one of the following clinical disease outcomes:

#### Non-Infected

Absence of positive culture and/or less than three positive Polymerase chain reaction (PCR) results for *B. pertussis* on or after day 6 after the challenge, regardless of reporting symptoms.

#### Asymptomatic Infection

At least one positive culture and/or three or more positive PCR results for *B. pertussis* on or after day 6 after the challenge in the absence of a pertussis-related symptom profile.

#### Symptomatic Infection

At least one positive culture and/or three or more positive PCR results for *B. pertussis* on or after day 6 after the challenge, plus a pertussis-related symptom profile (defined as two or more solicited pertussis symptoms, at least one of which must be a respiratory symptom) as described by ElSherif and colleagues (6).

All participants received azithromycin no later than day 19 or within 24-48 hours from cough onset, whichever occurred first. For this study, sera collected one day prior to challenge and at 14, 28, 56, 180, and 365 days post-inoculation were included in avidity analysis. Sera were stored at −80 Celsius prior to testing. PCR amplification data for the pertussis toxin subunit 1 (*ptxs1*) gene by PCR (reported as cycle threshold [Ct] values), along with *B. pertussis* CFU counts from nasal samples collected daily during the inpatient period were included in the analyses.

For this study, 20 symptomatic and 10 asymptomatic participants were selected for avidity analyses (Figure 1).

**Figure 1:**
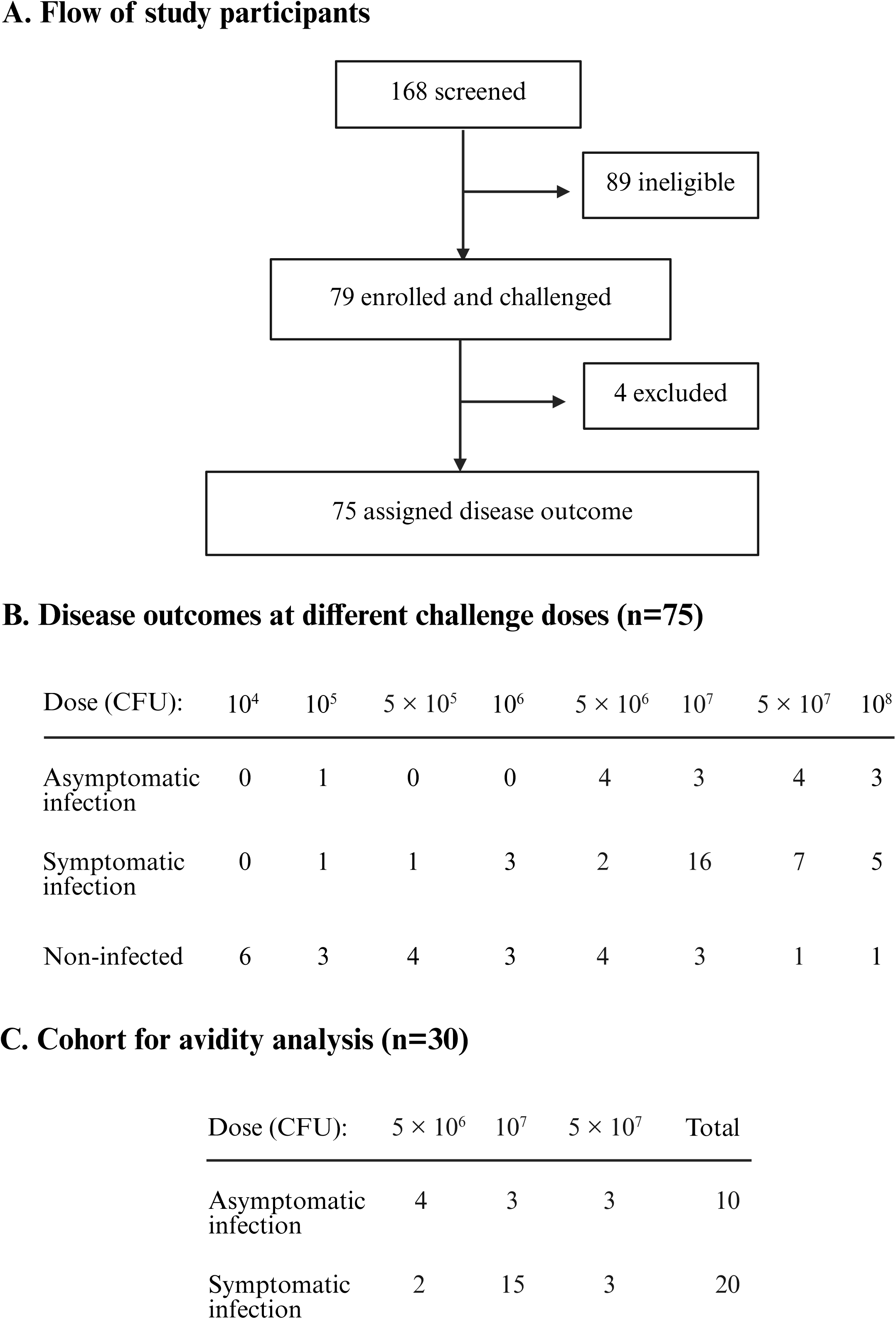
Scheme of the controlled human infection study and avidity cohort. (A) Controlled human infection study participants. (B) Participant distribution in different challenge dose groups. (C) Cohort of infected participants for avidity analyses.

### Measurement of total anti-PT IgG levels and anti-PT IgG avidity

Total anti-PT IgG levels and anti□PT IgG avidity were determined from sera using Enzyme-Linked Immunosorbent Assay (ELISA)-based elution (EUROIMMUN Medizinische Labordiagnostika, Lübeck, Germany) approach using a range of ammonium isothiocyanate (NH□SCN) at 0, 0·5, 1·0, 1·5, 2·0, or 3·0 Molar (SIGMA-ALDRICH, St. Louis, MO) as we published (7). Levels measured at 0 Molar of NH_4_SCN reflected total anti-PT IgG levels.

### Calculation of anti-PT IgG avidity indices

Different avidity indices were calculated as previously described (Supplementary Table□1) (7).

Relative avidity index (RAI) was calculated at each NH□SCN concentration as the percentage of anti□PT IgG remaining bound after treatment compared with untreated samples.

Fractional RAI, defined as the RAI achieved at a specific NH_4_SCN concentration, was calculated as the RAI at a specific concentration minus the RAI at the next higher NH_4_SCN concentration.

Total RAI (AU) reflected the weighted sum of the fractional RAIs across all NH□SCN concentrations and was calculated by applying a factor to each fractional RAI corresponding to the respective concentration of NH_4_SCN.

As indices involving RAI are relative measures, fractional and total absolute avidity levels were calculated. The fractional absolute avidity level of anti-PT IgG (IU/mL) reflected the level of anti-PT IgG that is still bound to the antigen at a specific NH_4_SCN concentration and calculated as the fractional RAI at a specific NH_4_SCN concentration multiplied by total anti-PT IgG. Fractional absolute avidities quantified at 0·5, 1, 1·5, 2, and 3 M of NH_4_SCN were classified as low-medium, medium, medium-high, high, and very high avidity antibodies, respectively. The levels of antibodies eluted (*i.e.* not bound to the plate) at the lowest NH_4_SCN concentration (0·5M) were classified as “low” avidity antibodies.

Total absolute avidity levels (AAU/mL) represented the weighted contribution of the fractional absolute avidities by applying a factor to each fractional absolute avidity level corresponding to the respective concentration of NH_4_SCN (Supplementary Table□1).

### Statistical analysis

Data normality was assessed using the Shapiro-Wilk test and visually inspected through Q-Q plots. Homoscedasticity and distribution randomness were evaluated using Levene’s test and the Runs test, respectively. As total anti-PT IgG and avidity indices data were not normally distributed, values were log_10_□transformed for statistical analyses, and geometric means with their 95% confidence intervals were calculated.

Total anti-PT IgG and avidity indices were compared between the pre-challenge day (day −1) and the time point when the levels peaked (to assess increase), between the peak and day 365 (to explore waning), and between day −1 and day 365 (to assess durability) using paired Welch’s t-test. Total anti-PT IgG and avidity indices were compared between symptomatic and asymptomatic participants using Welch’s t-test. Density estimates of total absolute avidity levels of anti-PT IgG for symptomatic and asymptomatic participants were performed using Gaussian kernels.

Principal component analysis (PCA) is a dimensionality-reduction technique that transforms a dataset with several variables into a smaller set of uncorrelated principal components that capture the maximum variance in the data. PCA analysis was performed using anti-PT IgG and avidity indices data. Unsupervised hierarchical clustering of the log_10_-transformed fractional absolute avidity levels of anti-PT IgG for all participants, standardized using z-score normalization, was used to visualize patterns and outliers without prior labelling of symptomatic or asymptomatic status.

Univariate linear regression model analyses were conducted to determine the association between covariates (clinical disease outcome [symptomatic *versus* asymptomatic], challenge dose, age, sex assigned at birth, weight, height, vaccine type used for primary vaccination series during the first 6 months of life [aP *versus* whole-cell pertussis (wP)], and time elapsed since the last pertussis vaccine dose) and the response variables (anti-PT IgG levels and avidity indices) at each time points. Because outcomes were log_10_-transformed, regression coefficients (β) were exponentiated and reported as fold-changes to facilitate interpretation on the original biological scale.

Covariates with p□values below 0·05 in univariate analyses were included in multivariate linear regression model analyses to estimate adjusted effects. Collinearity among covariates included in univariate analyses was assessed using linear regression models prior to inclusion in multivariate regression models.

A p-value <0·05 was considered statistically significant for all analyses. Statistical analyses were performed using RStudio v. 2026·01·0+392. Figures, tables, and graphs were generated using RStudio, Microsoft Excel, and Microsoft PowerPoint.

## Results

### Total anti-PT IgG and avidity kinetics

In this cohort, anti□PT IgG, total RAI and total absolute avidity levels increased following intranasal *B. pertussis* infection in all participants (n=30), reached their highest levels at day 56, and then declined through day 365 after inoculation with the bacteria (Figure 2).

**Figure 2:**
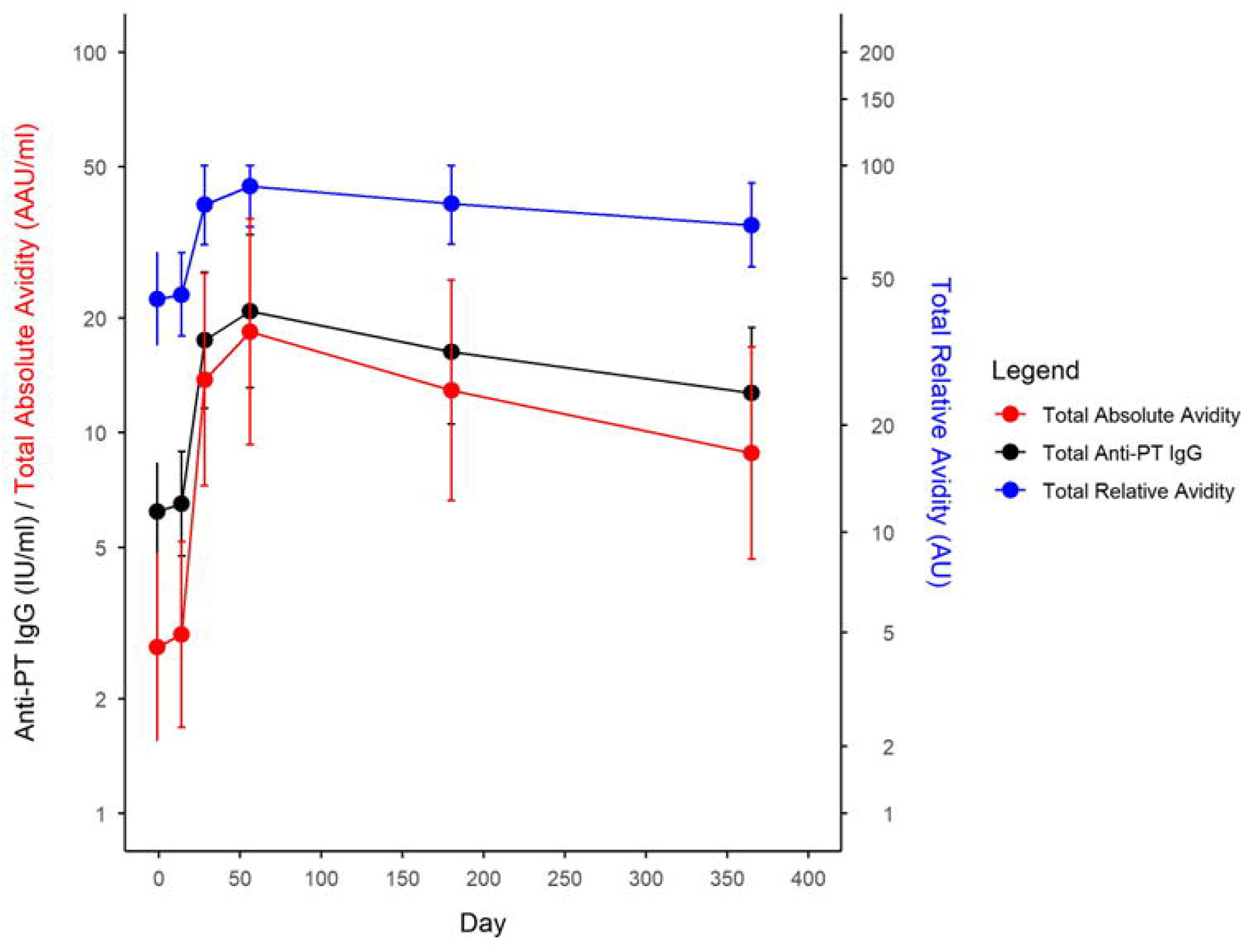
Kinetics of total anti□**pertussis toxin IgG (black), total relative avidity (blue), and total absolute avidity (red)**. Geometric means with 95% confidence intervals are shown as dots, and error bars, respectively. The x-axis represents one day prior (−1) and 14, 28, 56, 180, and 365 days post-inoculation with *Bordetella pertussis*.

### Increase in anti-PT IgG and avidity

Symptomatic participants exhibited statistically significant increases from pre-challenge to peak levels in total anti-PT IgG, total RAI, total absolute avidity, and all fractional avidity indices (Table S2), while asymptomatic participants had statistically significant increase only for low-medium avidity fraction.

### Waning of anti-PT IgG and avidity

In symptomatic participants, total anti-PT IgG, total RAI, total absolute avidity, and all fractional avidity categories (low-medium, medium-high, high, and very high) were significantly lower at day 365 compared with their respective peak timepoint (Table S2). Asymptomatic participants showed statistically significant decreases only in total RAI and low-medium avidity fractions at day 365 relative to peak timepoint (Table S2).

### Durability of anti-PT IgG and

At day 365 post-challenge, symptomatic participants maintained statistically higher levels of total anti-PT IgG, total RAI, total absolute avidity, and low-, medium-, and high-avidity antibodies compared with their pre-challenge baseline (Table S2). Among asymptomatic participants, total anti-PT IgG and all avidity indices at day 365 did not differ significantly from pre-challenge levels.

### Total anti-PT IgG, avidity and bacterial load by clinical disease outcome

Symptomatic participants developed significantly higher anti□PT IgG levels than those who remained asymptomatic at day 28, 56, 180 and 365 after challenge (Figure 3A, Table 1).

**Figure 3:**
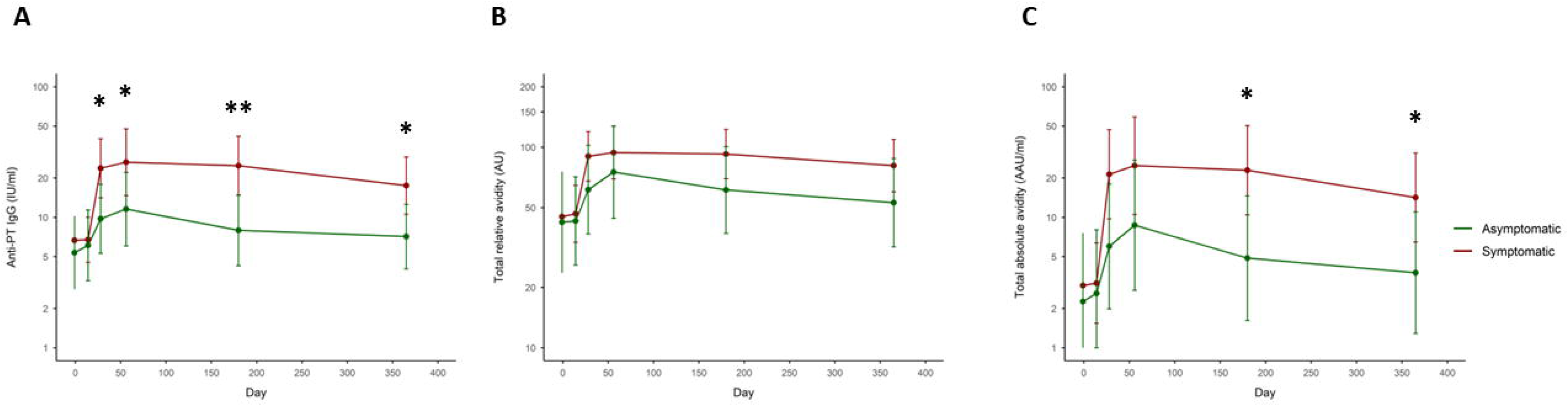
Kinetics of total anti□pertussis toxin IgG (A), total relative avidity (B), and total absolute avidity (C) in asymptomatic and symptomatic participants. Geometric means with 95% confidence intervals are shown as dots, and error bars, respectively. The x-axis represents one day prior (−1) and 14, 28, 56, 180, and 365 days post-inoculation with *Bordetella pertussis*. Statistical comparisons between asymptomatic and symptomatic groups at each time point were performed using unpaired Welch’s t test. *, *p*LJ<LJ0.05; **, *p*LJ<LJ0.01.

**Table 1:** Anti-PT IgG levels and avidity indices in asymptomatic and symptomatic participants.

|  | Asymptomatic (n=10) | Symptomatic (n=20) | p value (t-test) |
| --- | --- | --- | --- |
| **Visit 1 (Day -1)** |  |  |  |
| Total anti-PT IgG (IU/ml) | 5.35 [2.81, 10.21] | 6.67 [4.69, 9.49] | 0.52 |
| Total Relative Avidity (AU) | 42.25 [23.59, 75.66] | 45.06 [31.42, 64.62] | 0.84 |
| Total Absolute Avidity (AAU/ml) | 2.26 [0.68, 7.57] | 3.01 [1.51, 5.98] | 0.66 |
| **Fractional absolute anti-PT IgG levels per avidity range** |  |  |  |
| Low | 0.39 [0.09, 1.77] | 0.49 [0.21, 1.14] | 0.78 |
| Low-medium | 0.26 [0.07, 0.91] | 0.42 [0.18, 0.95] | 0.49 |
| Medium | 0.28 [0.07, 1.09] | 0.48 [0.20, 1.16] | 0.47 |
| Medium-high | 0.30 [0.07, 1.38] | 0.29 [0.13, 0.65] | 0.95 |
| High | 0.16 [0.06, 0.45] | 0.19 [0.11, 0.32] | 0.77 |
| Very high | 0.13 [0.07, 0.26] | 0.17 [0.12, 0.24] | 0.52 |
| **Visit 2 (Day 14)** |  |  |  |
| Total anti-PT IgG (IU/ml) | 6.09 [3.26, 11.38] | 6.73 [4.52, 10.03] | 0.77 |
| Total Relative Avidity (AU) | 42.91 [25.86, 71.20] | 46.56 [33.57, 64.59] | 0.77 |
| Total Absolute Avidity (AAU/ml) | 2.61 [0.85, 8.01] | 3.13 [1.54, 6.37] | 0.77 |
| **Fractional absolute anti-PT IgG levels per avidity range** |  |  |  |
| Low | 0.81 [0.16, 4.03] | 0.90 [0.32, 2.48] | 0.91 |
| Low-medium | 0.27 [0.08, 0.92] | 0.33 [0.16, 0.68] | 0.77 |
| Medium | 0.35 [0.09, 1.45] | 0.50 [0.20, 1.27] | 0.66 |
| Medium-high | 0.25 [0.07, 0.87] | 0.32 [0.14, 0.74] | 0.73 |
| High | 0.19 [0.07, 0.51] | 0.19 [0.11, 0.32] | 0.99 |
| Very high | 0.15 [0.08, 0.28] | 0.17 [0.11, 0.25] | 0.77 |
| **Visit 3 (Day 28)** |  |  |  |
| Total anti-PT IgG (IU/ml) | 9.75 [5.30, 17.90] | 23.75 [14.10, 40.02] | 0.02 |
| Total Relative Avidity (AU) | 61.44 [36.93, 102.21] | 90.03 [67.81, 119.52] | 0.17 |
| Total Absolute Avidity (AAU/ml) | 5.99 [1.99, 18.02] | 21.38 [9.72, 47.03] | 0.05 |
| **Fractional absolute anti-PT IgG levels per avidity range** |  |  |  |
| Low | 1.79 [0.48, 6.67] | 3.27 [1.75, 6.11] | 0.37 |
| Low-medium | 0.80 [0.22, 2.90] | 2.59 [1.06, 6.32] | 0.12 |
| Medium | 0.54 [0.17, 1.70] | 1.85 [0.83, 4.15] | 0.07 |
| Medium-high | 0.78 [0.18, 3.40] | 2.58 [1.01, 6.61] | 0.15 |
| High | 0.41 [0.12, 1.34] | 2.27 [0.74, 6.94] | 0.03 |
| Very high | 0.24 [0.13, 0.45] | 0.86 [0.40, 1.85] | 0.01 |
| **Visit 4 (Day 56)** |  |  |  |
| Total anti-PT IgG (IU/ml) | 11.56 [6.03, 22.15] | 26.43 [14.65, 47.67] | 0.05 |
| Total Relative Avidity (AU) | 75.19 [44.24, 127.77] | 94.10 [69.56, 127.31] | 0.42 |
| Total Absolute Avidity (AAU/ml) | 8.69 [2.76, 27.38] | 24.87 [10.52, 58.80] | 0.12 |
| **Fractional absolute anti-PT IgG levels per avidity range** |  |  |  |
| Low | 1.58 [0.44, 5.62] | 2.76 [1.41, 5.41] | 0.39 |
| Low-medium | 2.27 [0.63, 8.20] | 3.39 [1.51, 7.64] | 0.56 |
| Medium | 0.58 [0.17, 2.04] | 2.28 [0.92, 5.67] | 0.06 |
| Medium-high | 1·04 [0·20, 5·32] | 3·23 [1·17, 8·96] | 0·20 |
| High | 0·56 [0·12, 2·66] | 3·33 [1·10, 10·15] | 0·05 |
| Very high | 0·29 [0·15, 0·55] | 0·98 [0·43, 2·24] | 0·02 |
| **Visit 5 (Day 180)** |  |  |  |
| Total anti-PT IgG (IU/ml) | 7·95 [4·26, 14·82] | 24·83 [14·77, 41·76] | 0·005 |
| Total Relative Avidity (AU) | 61·21 [37·21, 100·69] | 92·46 [69·61, 122·81] | 0·13 |
| Total Absolute Avidity (AAU/ml) | 4·86 [1·62, 14·61] | 22·96 [10·44, 50·50] | 0·02 |
| **Fractional absolute anti-PT IgG levels per avidity range** |  |  |  |
| Low | 1·30 [0·39, 4·37] | 3·61 [1·86, 7·01] | 0·12 |
| Low-medium | 0·67 [0·17, 2·56] | 3·52 [1·63, 7·59] | 0·03 |
| Medium | 0·70 [0·17, 2·91] | 2·57 [1·05, 6·29] | 0·10 |
| Medium-high | 0·50 [0·12, 1·99] | 2·90 [1·09, 7·71] | 0·03 |
| High | 0·32 [0·10, 1·07] | 2·22 [0·73, 6·72] | 0·02 |
| Very high | 0·20 [0·11, 0·37] | 0·88 [0·41, 1·92] | 0·003 |
| **Visit 6 (Day 365)** |  |  |  |
| Total anti-PT IgG (IU/ml) | 7·12 [4·03, 12·58] | 17·52 [10·57, 29·05] | 0·02 |
| Total Relative Avidity (AU) | 52·88 [31·84, 87·82] | 80·95 [59·89, 109·41] | 0·13 |
| Total Absolute Avidity (AAU/ml) | 3·76 [1·29, 10·99] | 14·18 [6·47, 31·10] | 0·04 |
| **Fractional absolute anti-PT IgG levels per avidity range** |  |  |  |
| Low | 0·98 [0·25, 3·89] | 3·36 [1·49, 7·57] | 0·11 |
| Low-medium | 0·49 [0·14, 1·67] | 1·59 [0·78, 3·24] | 0·08 |
| Medium | 0·85 [0·19, 3·81] | 2·55 [1·05, 6·25] | 0·18 |
| Medium-high | 0·30 [0·09, 0·99] | 0·89 [0·38, 2·10] | 0·12 |
| High | 0·22 [0·09, 0·56] | 1·64 [0·53, 5·05] | 0·006 |
| Very high | 0·18 [0·10, 0·31] | 0·47 [0·26, 0·85] | 0·02 |

Geometric mean and 95% confidence intervals are shown for total anti-PT IgG, total relative avidity index, total absolute avidity, and fractional absolute avidity levels one day prior (-1) and 14, 28, 56, 180, and 365 days post-inoculation with *Bordetella pertussis*. Comparisons between groups were performed using Welch's two-sample t-test.

Total RAI levels did not differ significantly between symptomatic and asymptomatic participants at any time point (Figure 3B, Table 1). Total absolute avidity levels were statistically significantly higher in symptomatic *versus* asymptomatic participants at day 28, 180 and 365 after challenge (Figure 3C, Table 1).

Symptomatic participants had statistically significantly higher levels of high- and very high-avidity anti-PT IgG compared with asymptomatic participants at day 28, 56, 180 and 365 after challenge (Figure 4, Table 1). Symptomatic participants had also statistically significantly higher levels of anti-PT IgG with low-medium and medium-high avidity than asymptomatic participants on day 180 after challenge, and higher levels of medium avidity antibodies at 56 days post-challenge (Figure 4, Table 1).

**Figure 4:**
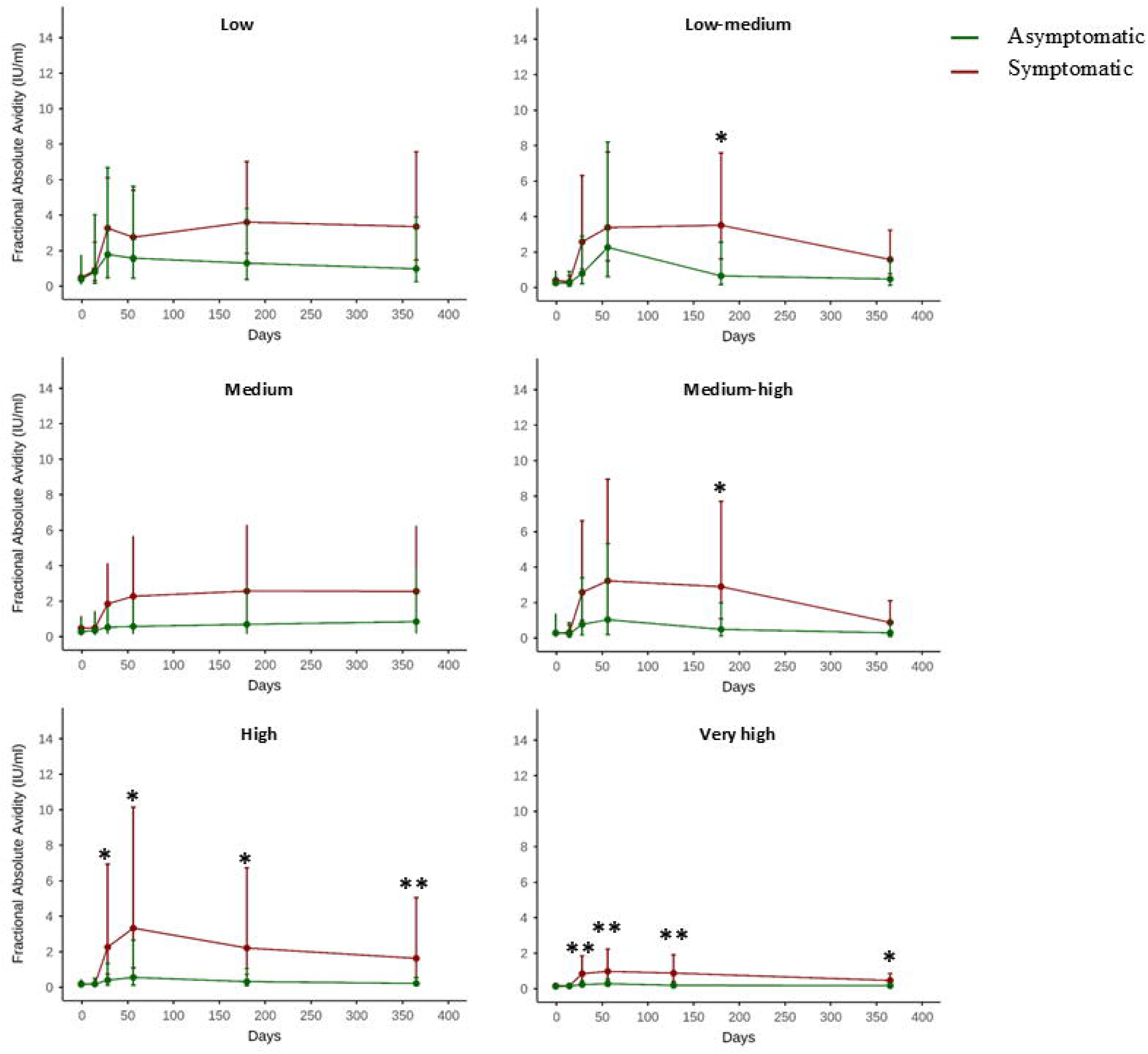
Kinetics of fractional absolute avidities. Fractional absolute avidity indices of anti-pertussis toxin IgG of the asymptomatic (green) and symptomatic (red) participants one day prior (−1) and 14, 28, 56, 180, and 365 days post-inoculation with *Bordetella pertussis*. Geometric mean and 95% confidence intervals of the geometric mean are shown as dots, and error bars, respectively. Statistical comparisons between asymptomatic and symptomatic groups at each time point were performed using unpaired Welch’s t test. *, *p*LJ<LJ0.05; **, *p*LJ<LJ0.01.

To explore whether avidity maturation was influenced by duration of antigenic exposure or levels of antigen gene expression at the mucosal level, we compared *B. pertussis* CFU counts from the nasal samples between groups. Symptomatic participants had significantly higher CFU counts on days 3, 7, and 13 post-challenge (Figure S1A), and statistically significantly higher *ptxs1* levels (reflected by lower Ct values) on day 7, 10 and 12 post-challenge (Figure S1B). CFU and *ptxs1* levels did not differ significantly between groups on the remaining days.

### Distribution of anti-PT IgG avidity by clinical disease outcome

To initially explore whether symptomatic and asymptomatic participants differed by avidity, density estimates were generated (Figure 5). Starting at day 28 post-inoculation, symptomatic participants demonstrated a right-shifted distribution, indicating a greater number of individuals with higher avidity antibodies compared with asymptomatic participants from this time point onward.

**Figure 5:**
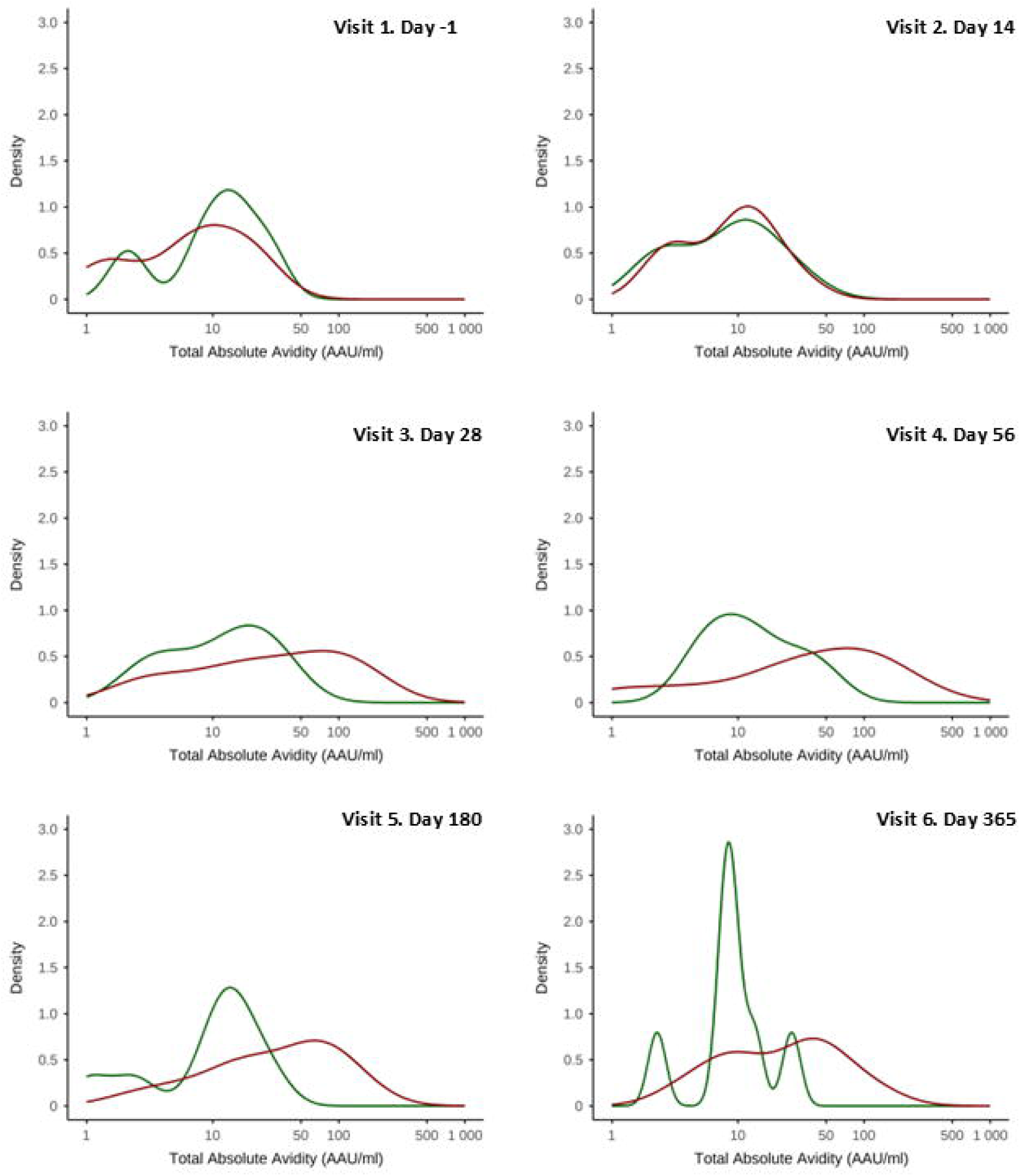
Distribution of total absolute avidities. Distribution of total absolute avidities of anti-pertussis toxin (PT) IgG of asymptomatic (green) and symptomatic (red) participants one day prior (−1) and 14, 28, 56, 180, and 365 days post-inoculation with *Bordetella pertussis*.

### Separation of symptomatic and asymptomatic participants by anti-PT IgG avidity

To further assess explore group separation based on avidity characteristics, PCA was conducted (Figure 6). Symptomatic and asymptomatic participants did not separate at baseline or on day 14. From day 28 onward, the symptomatic group showed a leftward displacement in principal component space.

**Figure 6:**
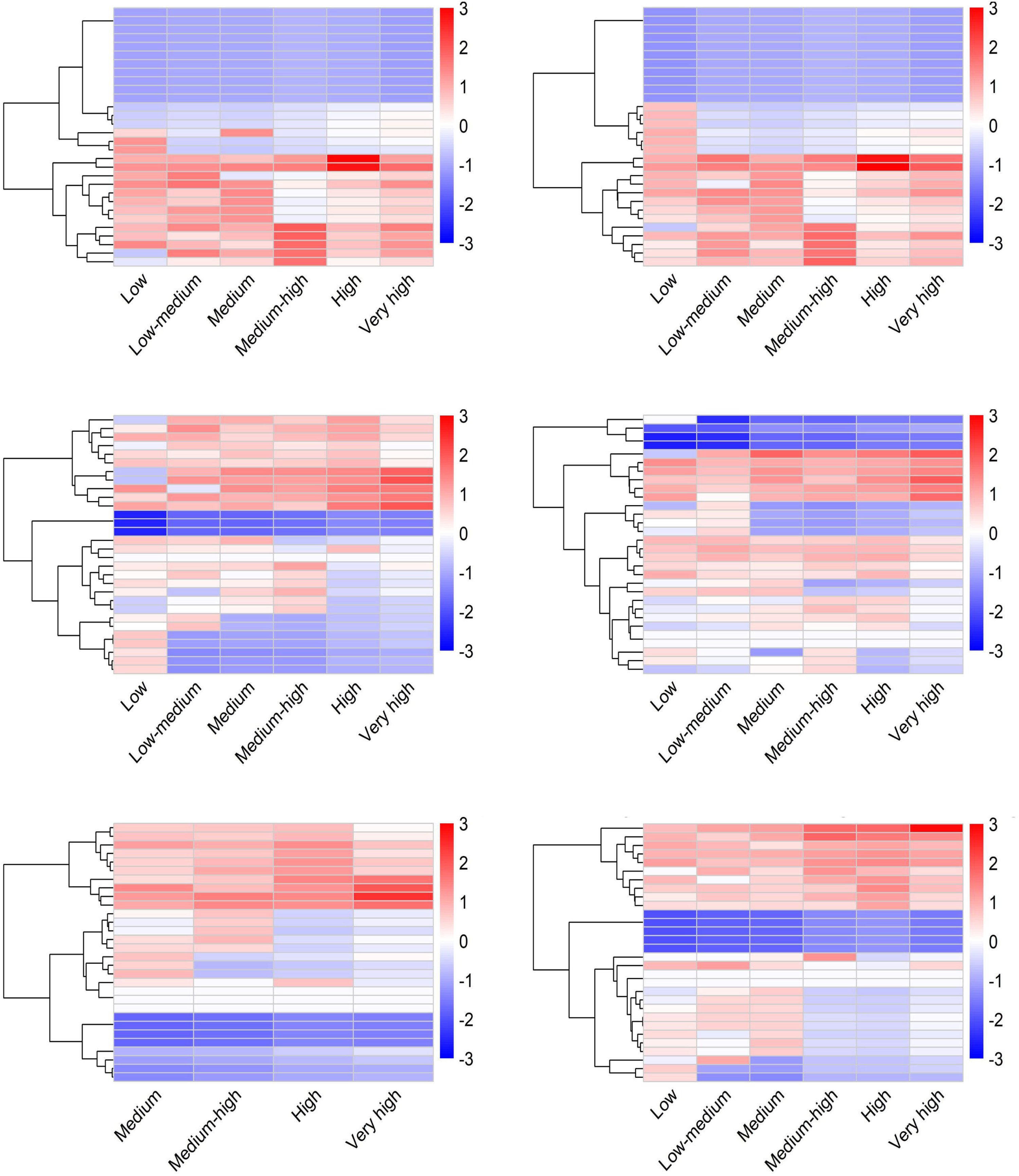
Heat map analyses of fractional absolute avidities. Unsupervised clustering of fractional anti-pertussis toxin (PT) IgG avidity one day prior (−1) and 14, 28, 56, 180, and 365 days post-inoculation with *Bordetella pertussis*. Heatmaps display z-score–standardized fractional absolute avidity indices of anti-PT IgG from symptomatic (red) and asymptomatic (green) participants.

### Clustering of symptomatic and asymptomatic participants by anti-PT IgG avidity

To evaluate clustering patterns of fractional absolute avidity values over time in symptomatic and asymptomatic participants, unsupervised hierarchical clustering was performed (Figure 7). Fractional absolute avidities did not clearly cluster symptomatic and asymptomatic participants before challenge and on day 14 after the inoculation. In contrast, more distinct clusters emerged on day 28 onwards, with symptomatic participants exhibiting higher levels of higher avidity antibodies relative to asymptomatic participants.

**Figure 7:**
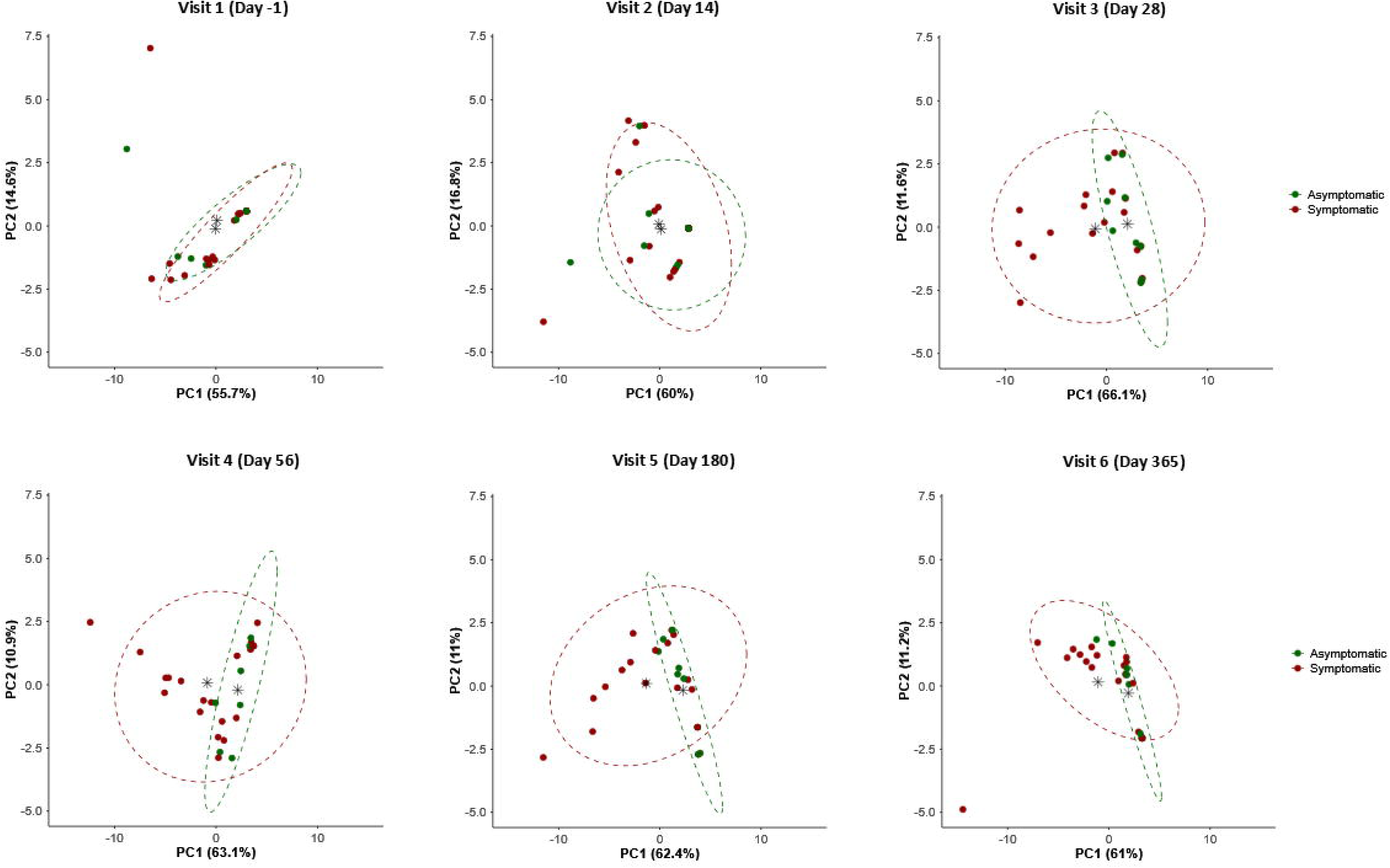
Principal component analyses of anti-PT IgG responses across study time points. Principal component analyses were performed using anti-pertussis toxin IgG, fractional relative avidity, and fractional absolute avidity data obtained from sera collected one day prior to inoculation (day –1) and at 14, 28, 56, 180, and 365 days following *Bordetella pertussis* challenge. Ellipsoids represent the distribution of asymptomatic (green) and symptomatic (red) participants, enclosing most data points from each group and illustrating their dispersion along the principal components. The centroid for each group, indicated by an asterisk, denotes the mean position of all samples from that group within the principal component space.

### Association between covariates and total anti-PT IgG, total RAI and total absolute avidity

#### Univariate analysis

There was no association between challenge dose, sex assigned at birth, height, or weight, and total anti-PT IgG, total RAI or total absolute avidity (Table 2). Symptomatic infection was associated with higher anti-PT IgG levels (at days 28, 180, and 365 post-inoculation), and total absolute avidity (at days 180 and 365). In contrast, longer time since the last pertussis vaccine was associated with lower anti-PT IgG concentrations at all time points, lower total RAI from day 14 onward, and lower total absolute avidity at all time points.

**Table 2.**
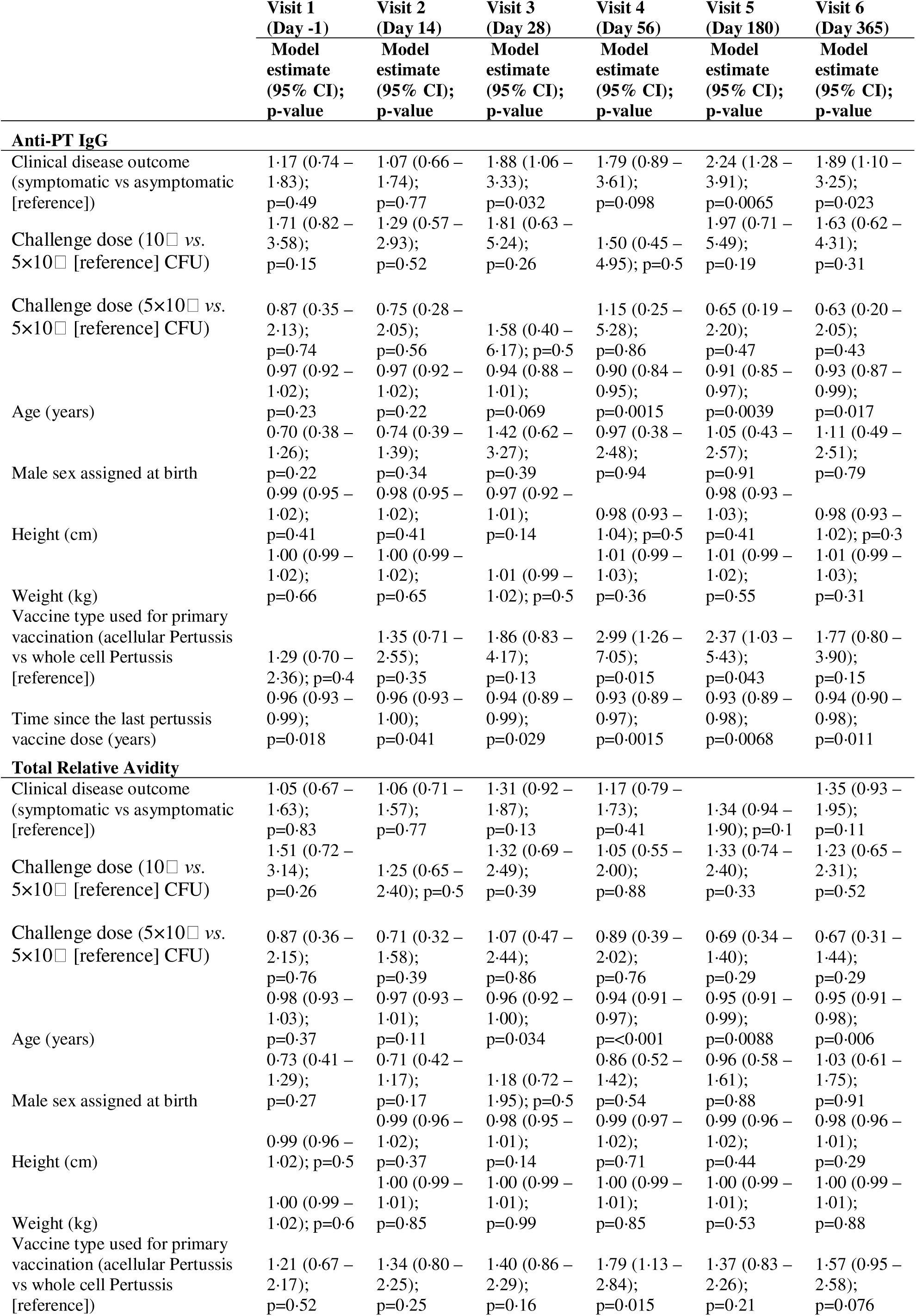
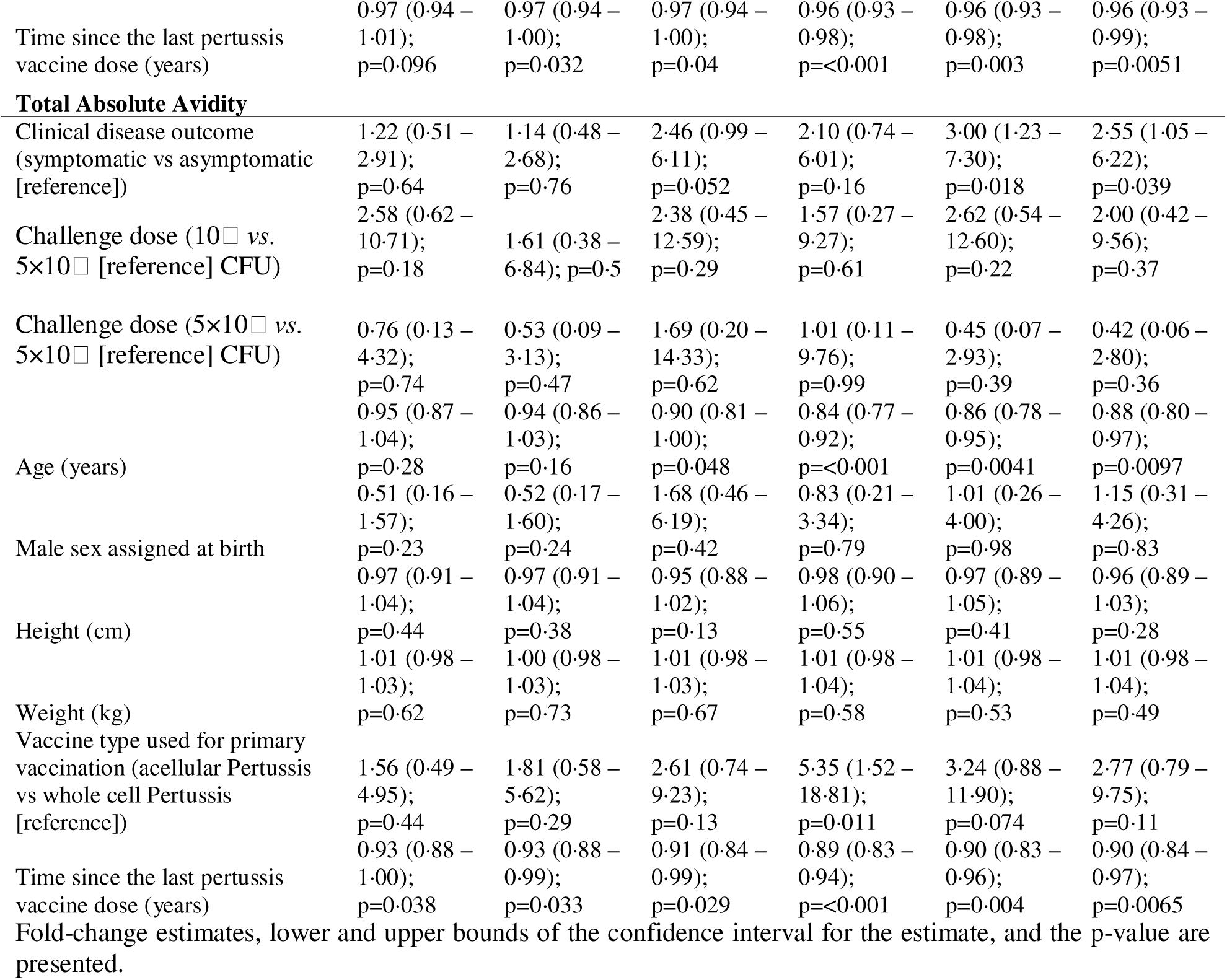
Univariate analyses of covariates with anti-PT IgG levels and avidity before challenge with *Bordetella pertussis* and different time point post challenge.

Additionally, older participants exhibited lower anti-PT IgG levels from day 56 onward, lower total relative avidity from day 28 onward, and lower total absolute avidity from day 56 onward. Priming with aP vaccines in infancy was associated with higher anti-PT IgG and total absolute avidity than with wP at day 56 and 180 post-challenge.

Age, vaccine type used for primary vaccination, and time since last pertussis vaccine dose were statistically significantly associated (co-correlated) (Table S3). Therefore, age was selected as the covariate to include in multivariate analyses.

#### Multivariate analysis

After adjusting for age, symptomatic infection remained associated with higher anti-PT IgG levels at days 180 and 365 post-challenge. Symptomatic infection was also associated with higher total absolute avidity at day 180 (Table 3).

**Table 3:** Multivariate analyses of covariates with anti-PT IgG levels and avidity before challenge with *Bordetella pertussis* and different time point post challenge.

|  | Day -1 | Day 14 | Day 28 | Day 56 | Day 180 | Day 365 |
| --- | --- | --- | --- | --- | --- | --- |
|  | Model estimate (95% CI); p-value | Model estimate (95% CI); p-value | Model estimate (95% CI); p-value | Model estimate (95% CI); p-value | Model estimate (95% CI); p-value | Model estimate (95% CI); p-value |
| **Anti-PT IgG** |  |  |  |  |  |  |
| Clinical disease outcome (symptomatic vs asymptomatic [reference]) | 1.13 (0.71–1.77); p=0.6 | 1.03 (0.63–1.67); p=0.9 | 1.72 (0.97–3.04); p=0.064 | 1.39 (0.74–2.61); p=0.29 | 2.02 (1.25–3.28); p=0.006 | 1.72 (1.04–2.87); p=0.037 |
| Age (years) | 0.97 (0.92–1.02); p=0.27 | 0.97 (0.92–1.02); p=0.24 | 0.95 (0.89–1.02); p=0.14 | 0.90 (0.85–0.97); p=0.0042 | 0.92 (0.87–0.97); p=0.0036 | 0.94 (0.88–0.99); p=0.028 |
| **Total relative avidity** |  |  |  |  |  |  |
| Clinical disease outcome (symptomatic vs asymptomatic [reference]) | 1.02 (0.65–1.59); p=0.94 | 1.01 (0.69–1.49); p=0.94 | 1.22 (0.86–1.73); p=0.25 | 1.00 (0.72–1.39); p=1 | 1.27 (0.92–1.74); p=0.14 | 1.26 (0.90–1.75); p=0.17 |
| Age (years) | 0.98 (0.93–1.03); p=0.39 | 0.97 (0.93–1.01); p=0.12 | 0.96 (0.92–1.00); p=0.061 | 0.94 (0.91–0.97); p=0.0011 | 0.95 (0.92–0.99); p=0.013 | 0.95 (0.91–0.99); p=0.01 |
| **Total absolute avidity** |  |  |  |  |  |  |
| Clinical disease outcome (symptomatic vs asymptomatic [reference]) | 1.14 (0.48–2.75); p=0.75 | 1.04 (0.44–2.45); p=0.92 | 2.10 (0.85–5.15); p=0.1 | 1.39 (0.56–3.45); p=0.46 | 2.56 (1.18–5.57); p=0.02 | 2.17 (0.96–4.91); p=0.062 |
| Age (years) | 0.95 (0.86–1.05); p=0.31 | 0.94 (0.85–1.03); p=0.17 | 0.92 (0.82–1.02); p=0.094 | 0.85 (0.77–0.93); p=0.0016 | 0.87 (0.80–0.96); p=0.0048 | 0.89 (0.81–0.98); p=0.016 |

Fold-change estimates, lower and upper bounds of the confidence interval for the estimate, and the p-value are presented.

#### Association between covariates and clinical disease outcome

At univariate analysis, clinical disease outcome was not associated with age, sex assigned at birth, height, weight, primary vaccination type (aP versus wP), or time since the last pertussis vaccine dose (Table 4). These findings support that the observed differences in antibody quantity and avidity were associated with clinical outcome rather than with demographic or vaccination background factors.

**Table 4:** Univariate analyses of covariates with clinical disease outcome.

|  | **Model estimate (95% CI); p-value** |
| --- | --- |
| Age (years) | -0.052 (-0.18 – 0.07); p=0.42 |
| Male sex assigned at birth | 0.41 (-1.12 – 2.01); p=0.61 |
| Height (cm) | -0.01 (-0.11 – 0.08), p=0.79 |
| Weight (kg) | -0.004 (-0.04 – 0.03); p=0.82 |
| Vaccine type used for primary vaccination (acellular Pertussis vs whole cell Pertussis [reference]) | 0.85 (-0.71 – 2.59); p=0.30 |
| Time since the last pertussis vaccine dose (years) | -0.044 (-0.14 – 0.05); p=0.33 |
| 453 Fold-change estimates, lower and upper bounds of the confidence interval for the estimate, and the p-value are |  |
| 454 presented. |  |

### Association between covariates and fractional absolute avidity

As total absolute avidity was associated with clinical disease outcome in multivariate analysis at 180 days after challenge (Table 3), associations between covariates (including clinical disease outcome) and different fractional absolute avidities were examined.

In univariate analysis, challenge dose, sex assigned at birth, height, and weight were not associated with fractional absolute avidities at any time point (Tables S4-S9). However, clinical disease outcome, age, primary vaccination, and time since last vaccination were associated with various fractional absolute avidity levels on day 28 onwards (Tables S4-S9). Given that three covariates (age, primary vaccination, and time since last vaccination) were co-correlated, only one of them (age) was included in the multivariate analyses. In multivariate analysis adjusted for age, symptomatic infection was associated with higher levels of high and very high avidity anti-PT IgG at day 180 and 365 post-inoculation, as well as low-medium, medium-high avidity anti-PT IgG on day 180 (Figure 8).

**Figure 8.**
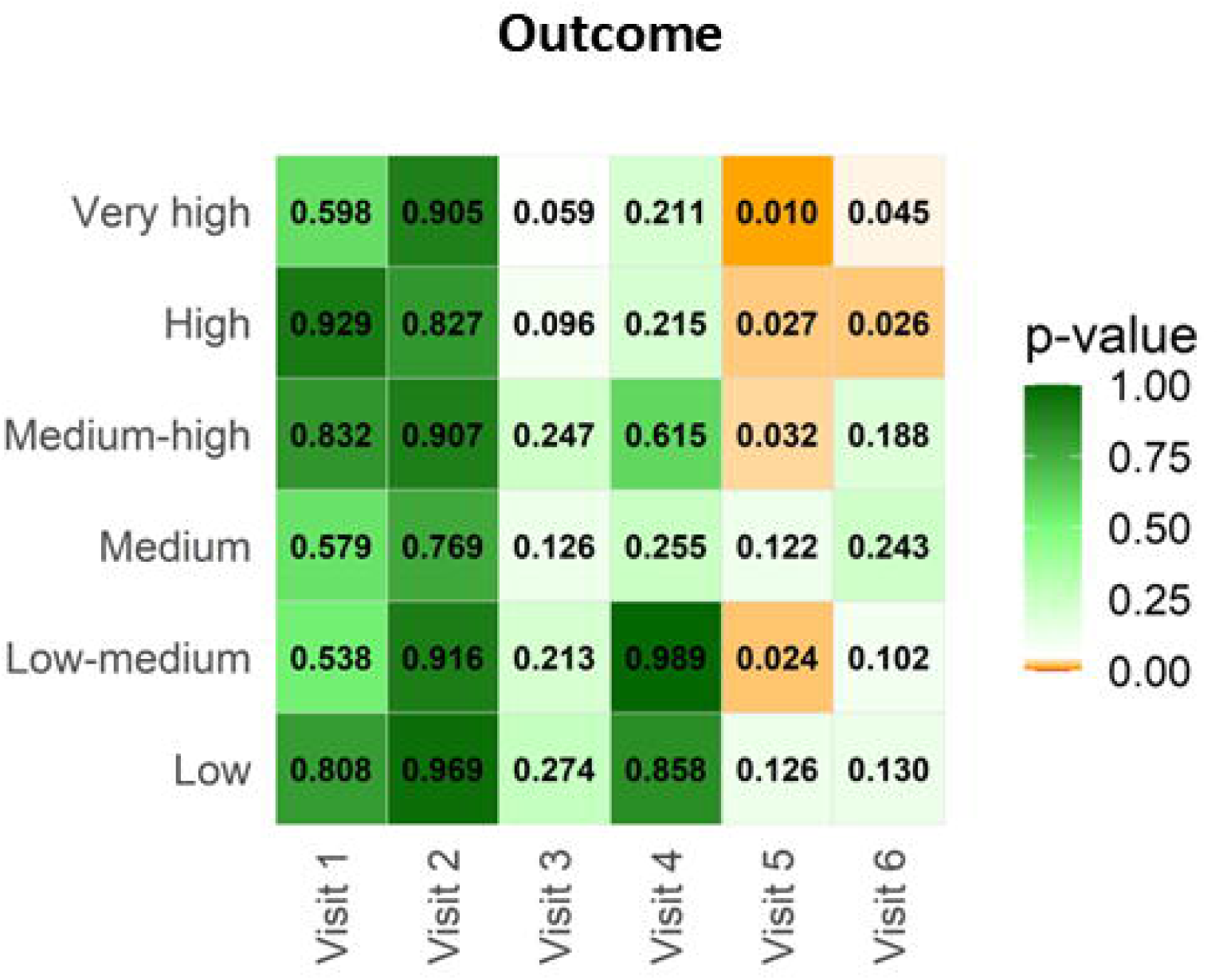
Multivariate analyses of fractional absolute avidity indices and clinical disease outcome. P-values from multivariate linear regression models evaluating the association between fractional absolute avidity indices (low, low-medium, medium, medium-high, high, and very high) and clinical disease outcome are shown for samples collected one day prior to inoculation (day –1) and at 14, 28, 56, 180, and 365 days following *Bordetella pertussis* challenge.

## Discussion

We show that avidity maturation is associated with symptomatic *B. pertussis* infection following intranasal challenge in a novel CHIM. These findings support that anti-PT IgG avidity may be a clinically relevant functional attribute of the antibody response to *B. pertussis* infection.

We found that symptomatic participants developed antibodies with higher avidities than those who remained asymptomatic and this was first noted 28 days after exposure to the bacteria, reached its peak at day 56 and persisted for one year after inoculation. The clinical relevance of these antibodies need further exploration and whether this immune response after recovery from infection might provide insight into immunological components associated with future protection, as expected in convalescence sera (8). Previous studies examining avidity maturation after pertussis infection are scarce. One study used a single concentration of diethylamine as a bond-breaking agent and reported increased anti-PT IgG avidity one month after confirmed infection in adolescents and adults (9). Another study reported higher avidity in aP-vaccinated adolescents and children compared with individuals with serologically diagnosed pertussis, although the interval between infection and sampling was unknown (4). These studies, conducted under natural-infection observational conditions, provided insights but illustrate limitations that can be overcome in CHIM trials. CHIM design enables precise definition of the challenge dose, use of well-defined solicited symptom criteria, and sample participants frequently at pre-specified time points—features that support more accurate comparison of symptomatic *versus* asymptomatic infection and their interplay with avidity maturation. CHIM studies are however limited by the fact that they must be conducted in healthy adults with previous vaccinations and episodes of infection. CHIM studies cannot be ethically performed in naïve young infants and children.

Although avidity maturation occurred in both outcome groups, asymptomatic individuals displayed more limited maturation. Asymptomatic infection could potentially lead to incomplete or short-lived affinity maturation, positioning asymptomatic individuals as a potential reservoir of susceptible hosts who may return to reinfection more rapidly than expected. Symptomatic infection elicited more pronounced avidity maturation, but even in these individuals, the progressive decline of high-avidity fractions could eventually increase susceptibility to reinfection or shift future infections toward an asymptomatic course. This interpretation is consistent with epidemiological and serological data showing recurrent circulation and reinfection across age groups despite widespread vaccination (24).

We observed that symptomatic participants mounted higher-avidity antibody responses regardless of the challenge dose administered, indicating that differences in avidity cannot be attributed solely to variation in inoculum size. Although circulating antigen could influence systemic affinity maturation, we did not assess antigenemia in this study. Indeed, systemic dissemination of PT can occur, as evidenced by its role in mediating characteristic systemic manifestations of pertussis, including leukocytosis and pulmonary hypertension (10). Another plausible explanation relates to antigen quantity and the duration of antigen availability at the mucosal site, which are established modulators of affinity maturation: very high antigen loads can reduce competitive selection in germinal centers, favoring lower-affinity B-cell clones (11), whereas sustained antigen availability supports positive selection of higher-affinity variants (12). However, comparable *B. pertussis* CFU counts and detection of *ptxS1* transcripts in nasal samples indicate that both symptomatic and asymptomatic participants encountered PT at the mucosal site. This suggests that differences in mucosal exposure alone are unlikely to explain the observed divergence in avidity.

Previous studies support the notion that antibody avidity contributes to functional protection and may decline even when total IgG levels remain detectable (13,14). However, previous studies examining durability of antibody avidity after pertussis infection are scarce. Hoving and colleagues found no significant change in anti-PT IgG avidity in PCR or serologically-confirmed pertussis symptomatic patients from the acute phase (median ∼ 3 weeks after diagnosis) to several years later (median 3.2 years after diagnosis), although avidity values were uniformly low (∼20%) and not stratified by binding strength (15). One explanation is that this may relate to the PT immunomodulatory activity, as this toxin interferes early in infection with innate immune recruitment and dendritic□cell trafficking —processes essential for germinal□center formation and affinity maturation (16–18)

Such disruption could truncate or distort germinal-center processes, contributing to the more rapid contraction of high-avidity fractions observed in our study. In this context, the decline of anti□PT antibodies described by others after infection (19) may largely reflect the persistence of lower□avidity antibodies, while functionally relevant higher□avidity fractions decline more rapidly than total anti-PT IgG measurements.

We show that anti□PT IgG avidity was independent of sex assigned at birth, challenge dose, height, and weight, but was associated with age, time since last pertussis vaccination, and vaccine type used for primary vaccination (wP *versus* aP). The association between anti-PT IgG avidity and age must be interpreted with cation due to collinearity among age, time since last vaccination, and primary vaccination vaccine type and thus we cannot conclude which one of covariates is mechanistically associated with avidity. Older participants were more likely to receive wP vaccines in infancy without an adolescent booster, whereas younger adults were more likely to receive aP vaccines and boosted during adolescence. Indeed, it was shown that adolescents who had been vaccinated within 5–8 years before *B. pertussis* infection had statistically significantly higher avidity in comparison to those who had been vaccinated 9–13 years before infection (4). Furthermore, the inverse association between age and high-avidity anti-PT IgG may suggest that repeated natural exposure (20), which is expected to have happened more in an older age group, may not sustain antibodies with high avidities, supporting the persistence of *B. pertussis* in the population.

This study has several strengths, including use of a well□characterized CHIM cohort with clearly defined clinical outcomes and frequent longitudinal sampling, enabling detailed characterization of avidity kinetics. We also applied a methodological approach for high□resolution avidity profiling (7,21,22). Our study still has some limitations. Firstly, the CHIM included only participants who had a baseline anti-PT IgG of < 20 IU/mL. It is important to measure avidity maturation following inoculation in a cohort of variable baseline anti-PT IgG levels. Secondly, disease progression was interrupted by the administration of Azithromycin, potentially affecting downstream immune maturation. Thirdly, CHIM dynamics may differ in magnitude or duration from natural community infections. Finally, while we focused on anti-PT IgG, immune response to other antigens may exhibit distinct avidity maturation patterns.

## Conclusions

Our study provides a novel longitudinal characterization of anti□PT antibody avidity in experimentally infected participants. Our findings identify antibody avidity as a functional marker associated with pertussis infection clinical disease endpoint and may point to its potential role in future work aimed at defining multicomponent correlates of immunity.

Additional studies should confirm these findings after natural pertussis exposures and outbreaks.

## Contribution

ME, KME, SH, BA contributed to the conceptualisation of the original CHIM and avidity study, clinical and laboratory operations. BA and CE conceptualized the analysis. CE, HO contributed to the data processing. CE and HO contributed to data analysis and visualization. All authors contributed to data interpretation. CE and BA drafted the paper. All authors critically reviewed and edited the paper. All authors approved the final decision for paper.

BA supervised all aspects of the work. Funding acquisition: BA (avidity study), SH (CHIM trial).

## Data sharing

Data request will be assessed upon request made to the corresponding author.

## Data Availability

Data available upon request

## Acknowledgements

We thank the trial staff, administrators, and laboratory technologists at the Canadian Centre for Vaccinology.

