## Supplementary file for "Avidity of anti-pertussis toxin antibodies is associated with symptomatic *Bordetella pertussis* infection in a novel controlled human infection model"

**Table S1: Calculation of relative avidity index, fractional relative avidity index, total relative avidity index and quantification of fractional and absolute avidity levels of anti-PT IgG.**

| NH_4_SCN concentration | **3 M** | **2 M** | **1**·**5 M** | **1 M** | **0**·**5 M** | **0 M** | **<0**·**5M **** |
| --- | --- | --- | --- | --- | --- | --- | --- |
| PT-IgG levels (IU/mL) | T_3M_ | T_2M_ | T_1.5M_ | T_1M_ | T_0.5M_ | T_0M_ | N/A |
| RAI^*^ (%) | RAI_3M_ = FRAI3M= | RAI_2M_ = T_2M_/T_0M_×100 | RAI_1.5M_ = T_1_·_5M_/T_0M_ ×100 | RAI_1_ = T_1M_/T_0M_ ×100 | RAI_0.5_ = T_0_·_5M_/T_0M_ ×100 | N/A | N/A |
| Fractional (F) RAI (%) | F RAI_3M_ = RAI_3M_ | F RAI_2M_ = RAI_2M_-RAI_3M_ | F RAI_1.5M_ = RAI_1·5M_-RAI_2M_ | F RAI_1M_ = RAI_1M_-RAI_1·5M_ | F RAI_0.5M_ = RAI_0·5M_-RAI_1M_ | N/A | F RAI_<0·5M_ = 100% - RAI_0·5M_ |
| Total RAI (AU) | (F RAI_3M_×3) + (F RAI_2M_×2) + (F RAI_1_·_5M_×1·5) + (F RAI_1M_×1) + (F RAI_0.5M_×0·5) + (F RAI_<0.5M_×0·25) | | | | | | |
| Fractional (F) absolute (abs) avidity levels (IU/mL) | F abs_3M_ =  F RAI_3M_×T_0M_ | F abs_2M_ =  F RAI_2M_×T_0M_ | F abs_1.5M_ =  F RAI_1·5M_×T_0M_ | F abs_1M_ =  F RAI_1M_×T_0M_ | F abs_0·5M_ =  F RAI_0·5M_×T_0M_ | N/A | F abs _<0·5M_ = FRAI_<0·5M_×T_0M_ |
| Total absolute avidity levels (AAU/mL) | (F abs_3M_×3) + (F abs_2M_×2) + (F abs_1·5M_×1.5) + (F abs_1M_×1) + (F abs_0·5M_×0.5) + (F abs_<0·5M_×0·25) | | | | | | |

**Abbreviations:** PT, pertussis toxin; IgG, immunoglobulin G; M, molar; N/A, not applicable; IU/mL, international unit/ml; T, total anti-PT IgG level; RAI, relative avidity index; F, fractional; AU, Avidity Unit; AAU/mL, Absolute Avidity Unit/mL; abs, absolute.

* Samples treated with 0·5M, 1M, 1·5M, 2M, 3M concentrations of NH_4_SCN and with optic density values lower than lower limit of quantitation in ELISA were assigned an arbitrary RAI value of 12·5%, 10%, 7·5%, 5%, 2·5% for each NH_4_SCN concentrations, respectively. The fractional absolute levels of antibodies quantified at 0·5 M, 1M, 1·5 M, 2M, and 3M of chaotrope were classified as low, low-medium, medium, medium-high, high, very high avidity antibodies, respectively. The levels of antibodies eluted by the lowest chaotrope concentration (<0·5 M) were classified as low avidity antibodies.

** This column includes the Fractional (F) RAI and Fractional (F) absolute (abs) avidity levels of PT-IgG antibodies eluted at the lowest NH_4_SCN concentration (Reproduced with minimal changes from Abu-Raya et al, Front. Immunol. 2019).

**Table S2.** Increase, waning, and durability of total anti‑PT IgG levels and avidity indices in symptomatic and asymptomatic participants.

|  |  | **Increase (p-value)** | **Waning (p-value)** | **Time of peak levels (days)** | **Durability (p-value)** |
| --- | --- | --- | --- | --- | --- |
| ***Symptomatic*** | ***Anti-PT IgG*** | <0·001 | 0·009 | 56 | 0·002 |
|  | ***Total relative avidity*** | <0·001 | <0·001 | 56 | 0·003 |
|  | ***Total absolute avidity*** | 0·002 | 0·008 | 56 | 0·005 |
|  | ***Fractional absolute avidity*** |  |  |  |  |
|  | Low | 0·004 | 0·462 | 28 | <0·001 |
|  | Low-medium | 0·005 | 0·021 | 28 | 0·054 |
|  | Medium | 0·007 | 0·941 | 180 | 0·003 |
|  | Medium-high | <0·001 | <0·001 | 56 | 0·188 |
|  | High | 0·004 | 0·024 | 56 | 0·004 |
|  | Very high | 0·009 | 0·025 | 56 | 0·089 |
| ***Asymptomatic*** | ***Anti-PT IgG*** | 0·096 | 0·108 |  | 0·367 |
|  | ***Total relative avidity*** | 0·1 | 0·029 |  | 0·343 |
|  | ***Total absolute avidity*** | 0·131 | 0·106 |  | 0·535 |
|  | ***Fractional absolute avidity*** |  |  |  |  |
|  | Low | 0·082 | 0·128 | 28 | 0·556 |
|  | Low-medium | 0·014 | 0·011 | 56 | 0·207 |
|  | Medium | 0·201 | N/A | 365 | 0·098 |
|  | Medium-high | 0·235 | 0·055 | 56 | 0·375 |
|  | High | 0·183 | 0·186 | 56 | 0·319 |
|  | Very high | 0·096 | 0·109 | 56 | 0·366 |

Statistical comparisons were performed between symptomatic and asymptomatic participants for the following immunological parameters: total anti‑PT IgG levels, total relative avidity, and total and fractional absolute avidities. The increase in immune responses was assessed by comparing the baseline levels measured at day −1 (one day prior to *Bordetella pertussis* challenge) with the corresponding values at the time points when the maximal levels were reached. Waning immunity was evaluated by comparing peak response with the level measured at the final time point (day 365). Durability was assessed by testing the difference between levels at day −1 and day 365. All comparisons were conducted using Welch’s paired t‑test, and statistically significant results are underlined (p < 0·05).

**Table S3. Correlation between co-variates.**

|  | Time since the last pertussis vaccine dose (years) | Vaccine type used for primary vaccination (acellular Pertussis vs whole cell Pertussis [reference]) | Clinical disease outcome (symptomatic vs asymptomatic [reference]) | Age (years) |
| --- | --- | --- | --- | --- |
| Vaccine type used for primary vaccination (acellular Pertussis vs whole cell Pertussis [reference]) | -0.464  (0.011) |  |  |  |
| Clinical disease outcome (symptomatic vs asymptomatic [reference]) | 0.308  (0.104) | -0.190  (0.314) |  |  |
| Age (years) | 0.651  (<0.001) | -0.842  (<0.001) | 0.156  (0.411) |  |
| Sex assigned at birth | 0.062  (0.750) | 0.144  (0.448) | 0.094  (0.619) | -0.093  (0.625) |

A Spearman correlation matrix was generated. Both rho correlation coefficients and p‑values are presented to illustrate the association strength and direction, and significance of these relationships, respectively. Statistically significant results are underlined (p < 0·05)

**Table S4.** Univariate analyses to determine association of clinical, vaccine and demographic factors with immunological properties of low avidity fractions before challenge with *Bordetella pertussis* and different time point post challenge.

|  | **Visit 1 (Day -1)** | **Visit 2 (Day 14)** | **Visit 3 (Day 28)** | **Visit 4 (Day 56)** | **Visit 5 (Day 180)** | **Visit 6 (Day 365)** |
| --- | --- | --- | --- | --- | --- | --- |
|  | **Model estimate (95% CI); p-value** | **Model estimate (95% CI); p-value** | **Model estimate (95% CI); p-value** | **Model estimate (95% CI); p-value** | **Model estimate (95% CI); p-value** | **Model estimate (95% CI); p-value** |
| **Anti-PT IgG** |  |  |  |  |  |  |
| Clinical disease outcome (symptomatic vs asymptomatic [reference]) | 1·02 (0·47 – 2·22); p=0·95 | 1·05 (0·68 – 1·61); p=0·83 | 1·00 (0·61 – 1·64); p=1 | 1·42 (0·80 – 2·52); p=0·22 | 1·44 (0·87 – 2·37); p=0·15 | 1·53 (0·96 – 2·43); p=0·072 |
| Challenge dose (10⁷ *vs.* 5×10⁶ [reference] CFU) | 1·51 (0·39 – 5·89); p=0·53 | 1·46 (0·71 – 3·02); p=0·29 | 1·44 (0·66 – 3·14); p=0·34 | 3·27 (1·42 – 7·50); p=0·0073 | 1·29 (0·56 – 2·97); p=0·54 | 1·62 (0·74 – 3·56); p=0·22 |
| Challenge dose (5×10⁷ *vs.* 5×10⁶ [reference] CFU) | 1·51 (0·21 – 10·65); p=0·66 | 0·97 (0·40 – 2·36); p=0·94 | 3·01 (1·08 – 8·39); p=0·037 | 3·28 (1·10 – 9·72); p=0·034 | 0·70 (0·22 – 2·30); p=0·54 | 0·97 (0·32 – 2·96); p=0·95 |
| Age (years) | 1·03 (0·94 – 1·13); p=0·5 | 0·97 (0·93 – 1·02); p=0·21 | 1·02 (0·97 – 1·08); p=0·4 | 0·95 (0·89 – 1·01); p=0·083 | 0·96 (0·90 – 1·02); p=0·17 | 0·99 (0·93 – 1·05); p=0·75 |
| Male sex assigned at birth | 1·10 (0·41 – 2·98); p=0·84 | 0·80 (0·45 – 1·41); p=0·43 | 1·17 (0·61 – 2·22); p=0·62 | 0·67 (0·32 – 1·41); p=0·28 | 0·91 (0·46 – 1·81); p=0·78 | 0·90 (0·47 – 1·73); p=0·74 |
| Height (cm) | 1·02 (0·97 – 1·07); p=0·45 | 0·98 (0·95 – 1·01); p=0·22 | 0·99 (0·95 – 1·03); p=0·61 | 0·98 (0·94 – 1·03); p=0·45 | 1·00 (0·96 – 1·04); p=0·93 | 1·00 (0·96 – 1·04); p=0·96 |
| Weight (kg) | 1·01 (0·99 – 1·03); p=0·39 | 1·00 (0·99 – 1·01); p=0·8 | 1·00 (0·98 – 1·01); p=0·61 | 0·99 (0·98 – 1·01); p=0·49 | 1·00 (0·98 – 1·01); p=0·85 | 1·01 (1·00 – 1·02); p=0·16 |
| Vaccine type used for primary vaccination (acellular Pertussis vs whole cell Pertussis [reference]) | 0·73 (0·28 – 1·90); p=0·5 | 1·45 (0·83 – 2·53); p=0·19 | 1·20 (0·63 – 2·28); p=0·57 | 1·94 (0·96 – 3·91); p=0·064 | 2·13 (1·17 – 3·87); p=0·016 | 1·02 (0·53 – 1·98); p=0·94 |
| Time since the last pertussis vaccine dose (years) | 0·96 (0·87 – 1·06); p=0·41 | 0·98 (0·95 – 1·01); p=0·16 | 1·01 (0·96 – 1·05); p=0·73 | 0·97 (0·93 – 1·02); p=0·19 | 0·99 (0·93 – 1·05); p=0·77 | 0·99 (0·93 – 1·06); p=0·82 |
| **Fractional relative avidity** | |  |  |  |  |  |
| Clinical disease outcome (symptomatic vs asymptomatic [reference]) | 1·00 (0·48 – 2·11); p=0·99 | 1·00 (0·43 – 2·34); p=1 | 0·82 (0·39 – 1·71); p=0·58 | 0·83 (0·44 – 1·56); p=0·54 | 0·92 (0·50 – 1·70); p=0·78 | 1·26 (0·69 – 2·31); p=0·44 |
| Challenge dose (10⁷ *vs.* 5×10⁶ [reference] CFU) | 2·42 (0·69 – 8·50); p=0·16 | 0·83 (0·19 – 3·53); p=0·79 | 1·10 (0·30 – 3·98); p=0·88 | 2·92 (1·13 – 7·53); p=0·028 | 0·68 (0·29 – 1·59); p=0·36 | 1·37 (0·54 – 3·48); p=0·5 |
| Challenge dose (5×10⁷ *vs.* 5×10⁶ [reference] CFU) | 1·44 (0·31 – 6·72); p=0·63 | 0·39 (0·07 – 2·33); p=0·29 | 1·26 (0·24 – 6·61); p=0·78 | 2·03 (0·61 – 6·81); p=0·24 | 0·18 (0·06 – 0·49); p=0·0018 | 0·38 (0·12 – 1·18); p=0·091 |
| Age (years) | 1·01 (0·93 – 1·09); p=0·86 | 0·96 (0·88 – 1·05); p=0·39 | 1·08 (1·00 – 1·18); p=0·05 | 0·98 (0·92 – 1·05); p=0·53 | 1·00 (0·93 – 1·08); p=0·94 | 0·98 (0·91 – 1·05); p=0·46 |
| Male sex assigned at birth | 0·73 (0·27 – 1·96); p=0·52 | 0·63 (0·20 – 1·92); p=0·4 | 1·84 (0·69 – 4·87); p=0·21 | 0·60 (0·27 – 1·33); p=0·2 | 0·60 (0·27 – 1·37); p=0·22 | 1·02 (0·45 – 2·35); p=0·96 |
| Height (cm) | 1·01 (0·95 – 1·07); p=0·72 | 0·99 (0·93 – 1·06); p=0·82 | 0·98 (0·92 – 1·03); p=0·38 | 0·99 (0·95 – 1·04); p=0·69 | 1·04 (0·99 – 1·09); p=0·15 | 0·99 (0·95 – 1·04); p=0·72 |
| Weight (kg) | 1·02 (1·00 – 1·04); p=0·068 | 1·00 (0·98 – 1·03); p=0·8 | 0·98 (0·96 – 1·00); p=0·068 | 0·98 (0·97 – 1·00); p=0·012 | 1·00 (0·98 – 1·02); p=0·84 | 1·00 (0·98 – 1·02); p=0·79 |
| Vaccine type used for primary vaccination (acellular Pertussis vs whole cell Pertussis [reference]) | 0·64 (0·24 – 1·71); p=0·36 | 0·91 (0·29 – 2·85); p=0·87 | 0·73 (0·27 – 1·97); p=0·52 | 1·23 (0·54 – 2·82); p=0·61 | 0·91 (0·39 – 2·15); p=0·83 | 1·12 (0·49 – 2·59); p=0·77 |
| Time since the last pertussis vaccine dose (years) | 0·96 (0·90 – 1·01); p=0·11 | 0·94 (0·88 – 0·99); p=0·033 | 1·05 (0·98 – 1·12); p=0·14 | 0·99 (0·94 – 1·04); p=0·58 | 0·98 (0·93 – 1·04); p=0·54 | 0·94 (0·90 – 0·98); p=0·0083 |
| **Fractional absolute avidity** | |  |  |  |  |  |
| Clinical disease outcome (symptomatic vs asymptomatic [reference]) | 1·17 (0·40 – 3·42); p=0·76 | 1·08 (0·31 – 3·69); p=0·9 | 1·53 (0·66 – 3·58); p=0·31 | 1·48 (0·61 – 3·61); p=0·37 | 2·06 (0·89 – 4·77); p=0·089 | 2·39 (0·88 – 6·47); p=0·085 |
| Challenge dose (10⁷ *vs.* 5×10⁶ [reference] CFU) | 4·15 (0·71 – 24·18); p=0·11 | 1·07 (0·13 – 8·74); p=0·95 | 1·99 (0·45 – 8·74); p=0·35 | 4·36 (1·14 – 16·78); p=0·033 | 1·33 (0·42 – 4·23); p=0·61 | 2·23 (0·44 – 11·15); p=0·32 |
| Challenge dose (5×10⁷ *vs.* 5×10⁶ [reference] CFU) | 1·25 (0·14 – 10·82); p=0·83 | 0·30 (0·02 – 3·87); p=0·34 | 1·99 (0·30 – 13·35); p=0·46 | 2·33 (0·42 – 12·98); p=0·32 | 0·11 (0·03 – 0·45); p=0·0034 | 0·24 (0·03 – 1·69); p=0·14 |
| Age (years) | 0·98 (0·87 – 1·10); p=0·7 | 0·93 (0·82 – 1·06); p=0·28 | 1·02 (0·92 – 1·13); p=0·73 | 0·88 (0·81 – 0·95); p=0·0022 | 0·91 (0·83 – 1·00); p=0·051 | 0·90 (0·81 – 1·01); p=0·08 |
| Male sex assigned at birth | 0·51 (0·12 – 2·07); p=0·33 | 0·46 (0·09 – 2·34); p=0·34 | 2·61 (0·86 – 7·91); p=0·087 | 0·58 (0·19 – 1·81); p=0·34 | 0·64 (0·19 – 2·14); p=0·45 | 1·14 (0·27 – 4·79); p=0·85 |
| Height (cm) | 1·00 (0·92 – 1·08); p=0·92 | 0·98 (0·89 – 1·08); p=0·63 | 0·94 (0·88 – 1·00); p=0·063 | 0·97 (0·91 – 1·04); p=0·4 | 1·01 (0·94 – 1·09); p=0·71 | 0·97 (0·89 – 1·05); p=0·42 |
| Weight (kg) | 1·02 (0·99 – 1·05); p=0·15 | 1·01 (0·97 – 1·04); p=0·73 | 0·99 (0·96 – 1·01); p=0·3 | 0·99 (0·97 – 1·01); p=0·35 | 1·01 (0·98 – 1·03); p=0·56 | 1·01 (0·98 – 1·04); p=0·67 |
| Vaccine type used for primary vaccination (acellular Pertussis vs whole cell Pertussis [reference]) | 0·82 (0·20 – 3·47); p=0·78 | 1·23 (0·23 – 6·42); p=0·8 | 1·36 (0·42 – 4·34); p=0·6 | 3·67 (1·27 – 10·57); p=0·018 | 2·17 (0·65 – 7·18); p=0·2 | 1·99 (0·48 – 8·25); p=0·33 |
| Time since the last pertussis vaccine dose (years) | 0·92 (0·85 – 0·99); p=0·034 | 0·90 (0·82 – 0·98); p=0·022 | 0·99 (0·91 – 1·07); p=0·77 | 0·91 (0·86 – 0·97); p=0·0029 | 0·92 (0·86 – 0·98); p=0·018 | 0·89 (0·82 – 0·96); p=0·0029 |

For each comparison, the fold‑change, lower and upper confidence interval bounds for the fold‑change, and the p‑value are reported.

**Table S5.** Univariate analyses to determine association of clinical, vaccinal and demographic factors with immunological properties of low-medium avidity fractions before challenge with *Bordetella pertussis* and different time point post challenge.

|  | **Visit 1 (Day -1)** | **Visit 2 (Day 14)** | **Visit 3 (Day 28)** | **Visit 4 (Day 56)** | **Visit 5 (Day 180)** | **Visit 6 (Day 365)** |
| --- | --- | --- | --- | --- | --- | --- |
|  | **Model estimate (95% CI); p-value** | **Model estimate (95% CI); p-value** | **Model estimate (95% CI); p-value** | **Model estimate (95% CI); p-value** | **Model estimate (95% CI); p-value** | **Model estimate (95% CI); p-value** |
| **Anti-PT IgG** |  |  |  |  |  |  |
| Clinical disease outcome (symptomatic vs asymptomatic [reference]) | 1·06 (0·69 – 1·64); p=0·77 | 0·90 (0·46 – 1·77); p=0·76 | 1·98 (1·01 – 3·85); p=0·046 | 1·86 (0·90 – 3·83); p=0·091 | 2·23 (1·24 – 4·01); p=0·0093 | 1·84 (1·08 – 3·15); p=0·027 |
| Challenge dose (10⁷ *vs.* 5×10⁶ [reference] CFU) | 1·60 (0·78 – 3·29); p=0·19 | 0·86 (0·28 – 2·60); p=0·77 | 2·04 (0·60 – 6·91); p=0·24 | 1·37 (0·39 – 4·76); p=0·61 | 2·05 (0·68 – 6·14); p=0·19 | 1·54 (0·57 – 4·15); p=0·37 |
| Challenge dose (5×10⁷ *vs.* 5×10⁶ [reference] CFU) | 0·95 (0·39 – 2·30); p=0·91 | 1·71 (0·32 – 9·15); p=0·51 | 1·62 (0·34 – 7·78); p=0·53 | 1·13 (0·23 – 5·54); p=0·88 | 0·83 (0·22 – 3·09); p=0·77 | 0·78 (0·24 – 2·56); p=0·66 |
| Age (years) | 0·98 (0·94 – 1·03); p=0·46 | 1·04 (0·96 – 1·14); p=0·33 | 0·91 (0·84 – 0·98); p=0·017 | 0·89 (0·83 – 0·95); p=0·0014 | 0·90 (0·85 – 0·97); p=0·004 | 0·92 (0·87 – 0·98); p=0·0075 |
| Male sex assigned at birth | 0·74 (0·42 – 1·31); p=0·29 | 1·61 (0·66 – 3·90); p=0·27 | 1·24 (0·47 – 3·28); p=0·65 | 0·90 (0·34 – 2·39); p=0·83 | 1·09 (0·43 – 2·75); p=0·85 | 1·14 (0·51 – 2·53); p=0·75 |
| Height (cm) | 0·99 (0·96 – 1·02); p=0·57 | 1·00 (0·95 – 1·05); p=0·98 | 0·96 (0·91 – 1·02); p=0·17 | 0·99 (0·93 – 1·04); p=0·65 | 0·97 (0·92 – 1·03); p=0·31 | 0·97 (0·93 – 1·02); p=0·19 |
| Weight (kg) | 1·00 (0·99 – 1·02); p=0·61 | 1·00 (0·99 – 1·02); p=0·74 | 1·01 (0·99 – 1·03); p=0·39 | 1·01 (0·99 – 1·03); p=0·31 | 1·01 (0·99 – 1·03); p=0·48 | 1·01 (0·99 – 1·02); p=0·32 |
| Vaccine type used for primary vaccination (acellular Pertussis vs whole cell Pertussis [reference]) | 1·31 (0·74 – 2·33); p=0·34 | 0·76 (0·32 – 1·82); p=0·52 | 2·15 (0·85 – 5·42); p=0·1 | 3·11 (1·28 – 7·58); p=0·014 | 2·45 (1·04 – 5·78); p=0·042 | 1·89 (0·88 – 4·07); p=0·099 |
| Time since the last pertussis vaccine dose (years) | 0·98 (0·94 – 1·01); p=0·16 | 1·02 (0·93 – 1·13); p=0·59 | 0·93 (0·88 – 0·98); p=0·011 | 0·92 (0·88 – 0·97); p=0·002 | 0·93 (0·89 – 0·98); p=0·0082 | 0·95 (0·91 – 0·99); p=0·022 |
| **Fractional relative avidity** |  |  |  |  |  |  |
| Clinical disease outcome (symptomatic vs asymptomatic [reference]) | 1·20 (0·66 – 2·18); p=0·53 | 1·07 (0·65 – 1·75); p=0·78 | 1·22 (0·63 – 2·39); p=0·54 | 0·74 (0·40 – 1·36); p=0·32 | 1·45 (0·81 – 2·60); p=0·21 | 1·21 (0·71 – 2·08); p=0·47 |
| Challenge dose (10⁷ *vs.* 5×10⁶ [reference] CFU) | 2·11 (0·79 – 5·63); p=0·13 | 1·39 (0·60 – 3·19); p=0·43 | 2·49 (0·87 – 7·11); p=0·085 | 1·36 (0·50 – 3·73); p=0·53 | 1·42 (0·52 – 3·86); p=0·47 | 1·70 (0·69 – 4·16); p=0·24 |
| Challenge dose (5×10⁷ *vs.* 5×10⁶ [reference] CFU) | 0·95 (0·29 – 3·17); p=0·94 | 0·77 (0·28 – 2·14); p=0·61 | 0·75 (0·19 – 2·88); p=0·66 | 1·24 (0·34 – 4·48); p=0·73 | 0·61 (0·19 – 2·02); p=0·41 | 0·86 (0·29 – 2·53); p=0·77 |
| Age (years) | 1·00 (0·94 – 1·07); p=0·95 | 0·96 (0·91 – 1·02); p=0·16 | 0·94 (0·87 – 1·01); p=0·097 | 0·99 (0·93 – 1·06); p=0·87 | 0·97 (0·90 – 1·03); p=0·31 | 0·95 (0·90 – 1·01); p=0·078 |
| Male sex assigned at birth | 0·67 (0·31 – 1·48); p=0·31 | 0·87 (0·45 – 1·68); p=0·68 | 1·23 (0·49 – 3·03); p=0·65 | 1·22 (0·56 – 2·69); p=0·6 | 0·82 (0·36 – 1·87); p=0·62 | 1·17 (0·56 – 2·44); p=0·66 |
| Height (cm) | 1·00 (0·95 – 1·05); p=0·95 | 0·97 (0·94 – 1·01); p=0·13 | 0·95 (0·90 – 1·00); p=0·034 | 0·98 (0·94 – 1·02); p=0·33 | 1·00 (0·95 – 1·05); p=0·97 | 0·97 (0·93 – 1·00); p=0·075 |
| Weight (kg) | 1·01 (0·99 – 1·03); p=0·33 | 1·00 (0·99 – 1·02); p=0·86 | 1·00 (0·98 – 1·02); p=0·84 | 1·00 (0·98 – 1·01); p=0·57 | 1·00 (0·98 – 1·02); p=0·95 | 1·00 (0·98 – 1·01); p=0·87 |
| Vaccine type used for primary vaccination (acellular Pertussis vs whole cell Pertussis [reference]) | 1·22 (0·55 – 2·74); p=0·61 | 1·50 (0·79 – 2·86); p=0·21 | 1·41 (0·57 – 3·47); p=0·44 | 1·04 (0·46 – 2·34); p=0·92 | 1·16 (0·50 – 2·68); p=0·72 | 1·49 (0·72 – 3·07); p=0·27 |
| Time since the last pertussis vaccine dose (years) | 0·97 (0·93 – 1·02); p=0·23 | 0·97 (0·94 – 1·01); p=0·17 | 0·95 (0·90 – 1·01); p=0·095 | 0·97 (0·93 – 1·02); p=0·21 | 0·95 (0·91 – 0·99); p=0·023 | 0·95 (0·91 – 0·98); p=0·0072 |
| **Fractional absolute avidity** |  |  |  |  |  |  |
| Clinical disease outcome (symptomatic vs asymptomatic [reference]) | 1·40 (0·52 – 3·77); p=0·49 | 1·15 (0·47 – 2·83); p=0·76 | 2·29 (0·81 – 6·49); p=0·11 | 1·33 (0·48 – 3·70); p=0·57 | 3·24 (1·25 – 8·42); p=0·018 | 2·30 (0·95 – 5·54); p=0·064 |
| Challenge dose (10⁷ *vs.* 5×10⁶ [reference] CFU) | 3·61 (0·72 – 18·04); p=0·11 | 1·79 (0·39 – 8·19); p=0·44 | 4·52 (0·78 – 26·28); p=0·09 | 2·04 (0·39 – 10·73); p=0·39 | 2·80 (0·53 – 14·93); p=0·22 | 2·77 (0·62 – 12·29); p=0·17 |
| Challenge dose (5×10⁷ *vs.* 5×10⁶ [reference] CFU) | 0·82 (0·11 – 5·92); p=0·84 | 0·58 (0·09 – 3·73); p=0·55 | 1·18 (0·12 – 11·35); p=0·88 | 1·42 (0·17 – 11·82); p=0·74 | 0·40 (0·05 – 2·93); p=0·35 | 0·54 (0·09 – 3·27); p=0·49 |
| Age (years) | 0·97 (0·87 – 1·09); p=0·62 | 0·93 (0·85 – 1·03); p=0·16 | 0·88 (0·78 – 0·99); p=0·036 | 0·89 (0·81 – 0·98); p=0·023 | 0·88 (0·78 – 0·98); p=0·021 | 0·88 (0·80 – 0·97); p=0·0099 |
| Male sex assigned at birth | 0·47 (0·13 – 1·72); p=0·24 | 0·65 (0·20 – 2·14); p=0·46 | 1·74 (0·41 – 7·48); p=0·44 | 1·18 (0·32 – 4·40); p=0·8 | 0·86 (0·20 – 3·75); p=0·84 | 1·30 (0·36 – 4·68); p=0·67 |
| Height (cm) | 0·98 (0·91 – 1·06); p=0·68 | 0·96 (0·89 – 1·03); p=0·21 | 0·92 (0·85 – 0·99); p=0·029 | 0·96 (0·89 – 1·03); p=0·28 | 0·98 (0·90 – 1·07); p=0·63 | 0·94 (0·88 – 1·01); p=0·089 |
| Weight (kg) | 1·01 (0·98 – 1·04); p=0·44 | 1·00 (0·98 – 1·03); p=0·73 | 1·00 (0·97 – 1·04); p=0·79 | 1·00 (0·98 – 1·03); p=0·75 | 1·01 (0·98 – 1·04); p=0·69 | 1·01 (0·98 – 1·03); p=0·58 |
| Vaccine type used for primary vaccination (acellular Pertussis vs whole cell Pertussis [reference]) | 1·58 (0·42 – 5·98); p=0·49 | 2·02 (0·62 – 6·62); p=0·23 | 2·62 (0·63 – 10·89); p=0·18 | 3·11 (0·88 – 11·05); p=0·077 | 2·74 (0·66 – 11·44); p=0·16 | 2·63 (0·77 – 9·00); p=0·12 |
| Time since the last pertussis vaccine dose (years) | 0·94 (0·87 – 1·01); p=0·071 | 0·94 (0·88 – 1·00); p=0·066 | 0·90 (0·82 – 0·98); p=0·02 | 0·90 (0·84 – 0·96); p=0·0028 | 0·89 (0·82 – 0·96); p=0·0033 | 0·89 (0·84 – 0·95); p=<0·001 |

For each comparison, the fold‑change, lower and upper confidence interval bounds for the fold‑change, and the p‑value are reported.

**Table S6.** Univariate analyses to determine association of clinical, vaccinal and demographic factors with immunological properties of medium avidity fractions before challenge with *Bordetella pertussis* and different time point post challenge.

|  | **Visit 1 (Day -1)** | **Visit 2 (Day 14)** | **Visit 3 (Day 28)** | **Visit 4 (Day 56)** | **Visit 5 (Day 180)** | **Visit 6 (Day 365)** |
| --- | --- | --- | --- | --- | --- | --- |
|  | **Model estimate (95% CI); p-value** | **Model estimate (95% CI); p-value** | **Model estimate (95% CI); p-value** | **Model estimate (95% CI); p-value** | **Model estimate (95% CI); p-value** | **Model estimate (95% CI); p-value** |
| **Anti-PT IgG** |  |  |  |  |  |  |
| Clinical disease outcome (symptomatic vs asymptomatic [reference]) | 1·01 (0·70 – 1·47); p=0·96 | 1·05 (0·72 – 1·53); p=0·77 | 1·92 (0·99 – 3·69); p=0·052 | 2·10 (0·98 – 4·54); p=0·057 | 2·03 (1·10 – 3·74); p=0·025 | 1·76 (1·03 – 3·02); p=0·041 |
| Challenge dose (10⁷ *vs.* 5×10⁶ [reference] CFU) | 1·35 (0·72 – 2·54); p=0·34 | 1·39 (0·73 – 2·63); p=0·3 | 1·59 (0·47 – 5·35); p=0·44 | 1·25 (0·32 – 4·80); p=0·74 | 2·14 (0·70 – 6·55); p=0·17 | 1·35 (0·50 – 3·69); p=0·54 |
| Challenge dose (5×10⁷ *vs.* 5×10⁶ [reference] CFU) | 0·95 (0·44 – 2·06); p=0·89 | 1·00 (0·46 – 2·20); p=0·99 | 1·62 (0·34 – 7·70); p=0·53 | 1·01 (0·18 – 5·65); p=0·99 | 1·02 (0·27 – 3·89); p=0·98 | 0·78 (0·23 – 2·61); p=0·67 |
| Age (years) | 0·98 (0·94 – 1·01); p=0·21 | 0·97 (0·94 – 1·01); p=0·2 | 0·92 (0·85 – 0·99); p=0·026 | 0·88 (0·82 – 0·94); p=<0·001 | 0·91 (0·85 – 0·97); p=0·0069 | 0·93 (0·87 – 0·99); p=0·016 |
| Male sex assigned at birth | 0·80 (0·49 – 1·30); p=0·36 | 0·81 (0·49 – 1·33); p=0·4 | 1·16 (0·45 – 2·99); p=0·76 | 0·82 (0·29 – 2·33); p=0·69 | 1·18 (0·47 – 2·98); p=0·72 | 1·06 (0·48 – 2·34); p=0·89 |
| Height (cm) | 0·99 (0·96 – 1·01); p=0·33 | 0·98 (0·96 – 1·01); p=0·26 | 0·98 (0·93 – 1·03); p=0·43 | 0·99 (0·93 – 1·05); p=0·77 | 0·97 (0·92 – 1·02); p=0·23 | 0·98 (0·94 – 1·02); p=0·36 |
| Weight (kg) | 1·00 (0·99 – 1·01); p=0·87 | 1·00 (0·99 – 1·01); p=0·84 | 1·01 (0·99 – 1·03); p=0·31 | 1·01 (0·99 – 1·03); p=0·31 | 1·01 (0·99 – 1·03); p=0·42 | 1·01 (0·99 – 1·03); p=0·29 |
| Vaccine type used for primary vaccination (acellular Pertussis vs whole cell Pertussis [reference]) | 1·35 (0·83 – 2·20); p=0·22 | 1·41 (0·86 – 2·29); p=0·16 | 2·06 (0·83 – 5·12); p=0·11 | 3·48 (1·34 – 9·04); p=0·012 | 2·30 (0·96 – 5·52); p=0·062 | 1·78 (0·82 – 3·83); p=0·14 |
| Time since the last pertussis vaccine dose (years) | 0·98 (0·95 – 1·01); p=0·18 | 0·98 (0·95 – 1·01); p=0·16 | 0·93 (0·88 – 0·99); p=0·02 | 0·92 (0·88 – 0·97); p=0·0032 | 0·94 (0·90 – 0·99); p=0·027 | 0·96 (0·92 – 1·00); p=0·077 |
| **Fractional relative avidity** |  |  |  |  |  |  |
| Clinical disease outcome (symptomatic vs asymptomatic [reference]) | 1·25 (0·63 – 2·51); p=0·51 | 1·19 (0·59 – 2·40); p=0·62 | 1·27 (0·80 – 2·03); p=0·3 | 1·47 (0·85 – 2·52); p=0·16 | 1·12 (0·62 – 2·04); p=0·69 | 1·15 (0·58 – 2·29); p=0·67 |
| Challenge dose (10⁷ *vs.* 5×10⁶ [reference] CFU) | 1·64 (0·51 – 5·25); p=0·39 | 2·01 (0·63 – 6·41); p=0·23 | 1·38 (0·61 – 3·12); p=0·43 | 0·99 (0·39 – 2·50); p=0·99 | 2·86 (1·13 – 7·21); p=0·028 | 1·17 (0·36 – 3·84); p=0·79 |
| Challenge dose (5×10⁷ *vs.* 5×10⁶ [reference] CFU) | 0·61 (0·15 – 2·52); p=0·48 | 0·73 (0·18 – 3·00); p=0·65 | 1·01 (0·35 – 2·89); p=0·98 | 0·81 (0·25 – 2·62); p=0·71 | 1·21 (0·40 – 3·66); p=0·72 | 0·74 (0·18 – 3·11); p=0·67 |
| Age (years) | 0·96 (0·89 – 1·03); p=0·24 | 0·97 (0·90 – 1·05); p=0·43 | 0·96 (0·91 – 1·01); p=0·096 | 0·94 (0·89 – 0·99); p=0·024 | 0·96 (0·90 – 1·03); p=0·21 | 0·94 (0·87 – 1·01); p=0·096 |
| Male sex assigned at birth | 0·78 (0·31 – 1·98); p=0·59 | 0·69 (0·27 – 1·74); p=0·42 | 1·03 (0·54 – 1·95); p=0·94 | 1·02 (0·50 – 2·11); p=0·95 | 1·17 (0·51 – 2·67); p=0·7 | 0·91 (0·36 – 2·31); p=0·84 |
| Height (cm) | 0·97 (0·92 – 1·03); p=0·3 | 0·97 (0·92 – 1·03); p=0·32 | 0·99 (0·96 – 1·03); p=0·62 | 0·98 (0·94 – 1·02); p=0·37 | 0·95 (0·91 – 1·00); p=0·037 | 0·97 (0·92 – 1·02); p=0·27 |
| Weight (kg) | 1·01 (0·99 – 1·03); p=0·29 | 1·01 (0·99 – 1·03); p=0·41 | 1·00 (0·99 – 1·02); p=0·63 | 1·01 (0·99 – 1·02); p=0·38 | 1·00 (0·99 – 1·02); p=0·69 | 1·00 (0·98 – 1·02); p=0·83 |
| Vaccine type used for primary vaccination (acellular Pertussis vs whole cell Pertussis [reference]) | 1·79 (0·72 – 4·47); p=0·2 | 1·58 (0·62 – 4·02); p=0·32 | 1·22 (0·64 – 2·32); p=0·53 | 1·60 (0·78 – 3·27); p=0·19 | 1·12 (0·49 – 2·58); p=0·78 | 1·36 (0·54 – 3·46); p=0·5 |
| Time since the last pertussis vaccine dose (years) | 0·96 (0·91 – 1·02); p=0·16 | 0·96 (0·91 – 1·02); p=0·17 | 0·95 (0·92 – 0·99); p=0·02 | 0·95 (0·92 – 0·99); p=0·01 | 0·96 (0·91 – 1·00); p=0·056 | 0·95 (0·90 – 1·00); p=0·055 |
| **Fractional absolute avidity** |  |  |  |  |  |  |
| Clinical disease outcome (symptomatic vs asymptomatic [reference]) | 1·46 (0·51 – 4·20); p=0·46 | 1·27 (0·42 – 3·89); p=0·66 | 2·39 (0·94 – 6·09); p=0·066 | 2·63 (0·86 – 8·02); p=0·086 | 2·51 (0·87 – 7·28); p=0·087 | 2·18 (0·73 – 6·50); p=0·15 |
| Challenge dose (10⁷ *vs.* 5×10⁶ [reference] CFU) | 2·81 (0·50 – 15·82); p=0·23 | 2·60 (0·41 – 16·57); p=0·3 | 2·50 (0·46 – 13·56); p=0·28 | 1·48 (0·22 – 10·18); p=0·68 | 5·63 (1·00 – 31·56); p=0·05 | 1·91 (0·28 – 12·83); p=0·49 |
| Challenge dose (5×10⁷ *vs.* 5×10⁶ [reference] CFU) | 0·53 (0·06 – 4·36); p=0·54 | 0·55 (0·06 – 5·26); p=0·59 | 1·60 (0·18 – 14·05); p=0·66 | 0·92 (0·08 – 10·77); p=0·95 | 0·78 (0·10 – 6·15); p=0·81 | 0·47 (0·05 – 4·67); p=0·5 |
| Age (years) | 0·93 (0·83 – 1·04); p=0·2 | 0·94 (0·83 – 1·06); p=0·3 | 0·90 (0·81 – 1·00); p=0·051 | 0·84 (0·76 – 0·93); p=0·0019 | 0·87 (0·77 – 0·98); p=0·021 | 0·87 (0·77 – 0·98); p=0·023 |
| Male sex assigned at birth | 0·54 (0·13 – 2·20); p=0·38 | 0·51 (0·12 – 2·21); p=0·35 | 1·46 (0·38 – 5·55); p=0·57 | 0·99 (0·22 – 4·47); p=0·99 | 1·23 (0·26 – 5·81); p=0·78 | 1·01 (0·22 – 4·76); p=0·99 |
| Height (cm) | 0·96 (0·88 – 1·04); p=0·3 | 0·96 (0·88 – 1·04); p=0·32 | 0·96 (0·89 – 1·03); p=0·24 | 0·96 (0·89 – 1·05); p=0·39 | 0·93 (0·85 – 1·02); p=0·12 | 0·95 (0·87 – 1·03); p=0·22 |
| Weight (kg) | 1·01 (0·98 – 1·04); p=0·38 | 1·01 (0·98 – 1·04); p=0·47 | 1·01 (0·98 – 1·04); p=0·51 | 1·02 (0·98 – 1·05); p=0·32 | 1·01 (0·98 – 1·04); p=0·58 | 1·01 (0·97 – 1·04); p=0·68 |
| Vaccine type used for primary vaccination (acellular Pertussis vs whole cell Pertussis [reference]) | 2·31 (0·57 – 9·30); p=0·23 | 2·13 (0·49 – 9·31); p=0·3 | 2·27 (0·61 – 8·37); p=0·21 | 4·78 (1·17 – 19·59); p=0·031 | 2·65 (0·58 – 12·10); p=0·2 | 2·41 (0·53 – 10·98); p=0·24 |
| Time since the last pertussis vaccine dose (years) | 0·92 (0·85 – 1·00); p=0·054 | 0·93 (0·85 – 1·01); p=0·08 | 0·90 (0·83 – 0·97); p=0·012 | 0·88 (0·82 – 0·95); p=0·001 | 0·89 (0·82 – 0·97); p=0·0091 | 0·90 (0·82 – 0·97); p=0·011 |

For each comparison, the fold‑change, lower and upper confidence interval bounds for the fold‑change, and the p‑value are reported.

**Table S7.** Univariate analyses to determine association of clinical, vaccinal and demographic factors with immunological properties of medium-high avidity fractions before challenge with *Bordetella pertussis* and different time point post challenge.

|  | **Visit 1 (Day -1)** | **Visit 2 (Day 14)** | **Visit 3 (Day 28)** | **Visit 4 (Day 56)** | **Visit 5 (Day 180)** | **Visit 6 (Day 365)** |
| --- | --- | --- | --- | --- | --- | --- |
|  | **Model estimate (95% CI); p-value** | **Model estimate (95% CI); p-value** | **Model estimate (95% CI); p-value** | **Model estimate (95% CI); p-value** | **Model estimate (95% CI); p-value** | **Model estimate (95% CI); p-value** |
| **Anti-PT IgG** |  |  |  |  |  |  |
| Clinical disease outcome (symptomatic vs asymptomatic [reference]) | 0·94 (0·68 – 1·29); p=0·68 | 1·03 (0·77 – 1·37); p=0·86 | 1·82 (0·96 – 3·44); p=0·067 | 2·03 (0·95 – 4·32); p=0·066 | 2·08 (1·15 – 3·74); p=0·017 | 1·74 (1·01 – 3·01); p=0·047 |
| Challenge dose (10⁷ *vs.* 5×10⁶ [reference] CFU) | 1·21 (0·69 – 2·10); p=0·5 | 1·16 (0·70 – 1·93); p=0·56 | 1·44 (0·44 – 4·66); p=0·53 | 1·17 (0·31 – 4·38); p=0·81 | 1·54 (0·50 – 4·78); p=0·44 | 1·23 (0·44 – 3·42); p=0·68 |
| Challenge dose (5×10⁷ *vs.* 5×10⁶ [reference] CFU) | 1·09 (0·55 – 2·16); p=0·79 | 1·11 (0·60 – 2·07); p=0·73 | 1·63 (0·36 – 7·39); p=0·51 | 1·04 (0·19 – 5·63); p=0·96 | 0·98 (0·25 – 3·80); p=0·98 | 0·80 (0·23 – 2·75); p=0·72 |
| Age (years) | 0·98 (0·95 – 1·02); p=0·32 | 0·98 (0·95 – 1·01); p=0·1 | 0·92 (0·86 – 0·99); p=0·025 | 0·88 (0·82 – 0·94); p=<0·001 | 0·91 (0·85 – 0·97); p=0·0046 | 0·93 (0·88 – 0·99); p=0·032 |
| Male sex assigned at birth | 0·80 (0·53 – 1·22); p=0·3 | 0·85 (0·58 – 1·25); p=0·4 | 1·10 (0·44 – 2·77); p=0·83 | 0·79 (0·28 – 2·20); p=0·63 | 1·13 (0·46 – 2·81); p=0·78 | 1·10 (0·49 – 2·45); p=0·81 |
| Height (cm) | 0·99 (0·97 – 1·02); p=0·53 | 0·99 (0·97 – 1·01); p=0·41 | 0·98 (0·93 – 1·03); p=0·48 | 1·00 (0·94 – 1·06); p=0·89 | 0·98 (0·93 – 1·03); p=0·44 | 0·99 (0·94 – 1·03); p=0·6 |
| Weight (kg) | 1·00 (0·99 – 1·01); p=0·45 | 1·00 (0·99 – 1·01); p=0·55 | 1·01 (0·99 – 1·03); p=0·36 | 1·01 (0·99 – 1·03); p=0·42 | 1·01 (0·99 – 1·03); p=0·49 | 1·01 (0·99 – 1·03); p=0·25 |
| Vaccine type used for primary vaccination (acellular Pertussis vs whole cell Pertussis [reference]) | 1·22 (0·80 – 1·86); p=0·35 | 1·37 (0·94 – 1·99); p=0·095 | 2·11 (0·88 – 5·04); p=0·092 | 3·54 (1·40 – 8·93); p=0·0094 | 2·47 (1·07 – 5·71); p=0·036 | 1·87 (0·86 – 4·03); p=0·11 |
| Time since the last pertussis vaccine dose (years) | 0·99 (0·97 – 1·02); p=0·45 | 0·99 (0·97 – 1·01); p=0·36 | 0·94 (0·89 – 1·00); p=0·044 | 0·93 (0·88 – 0·98); p=0·0086 | 0·95 (0·90 – 1·00); p=0·065 | 0·97 (0·93 – 1·02); p=0·24 |
| **Fractional relative avidity** |  |  |  |  |  |  |
| Clinical disease outcome (symptomatic vs asymptomatic [reference]) | 0·83 (0·42 – 1·65); p=0·58 | 1·10 (0·58 – 2·11); p=0·76 | 1·24 (0·63 – 2·48); p=0·52 | 1·24 (0·60 – 2·58); p=0·55 | 1·55 (0·79 – 3·05); p=0·19 | 1·14 (0·68 – 1·93); p=0·61 |
| Challenge dose (10⁷ *vs.* 5×10⁶ [reference] CFU) | 1·17 (0·35 – 3·94); p=0·8 | 1·27 (0·41 – 3·91); p=0·66 | 1·48 (0·45 – 4·91); p=0·5 | 0·88 (0·27 – 2·92); p=0·83 | 1·00 (0·30 – 3·39); p=1 | 0·93 (0·38 – 2·32); p=0·88 |
| Challenge dose (5×10⁷ *vs.* 5×10⁶ [reference] CFU) | 0·95 (0·21 – 4·19); p=0·94 | 0·77 (0·19 – 3·03); p=0·7 | 1·45 (0·31 – 6·75); p=0·62 | 0·63 (0·14 – 2·89); p=0·53 | 0·80 (0·19 – 3·43); p=0·75 | 0·66 (0·22 – 1·99); p=0·45 |
| Age (years) | 0·97 (0·90 – 1·05); p=0·48 | 0·95 (0·88 – 1·01); p=0·12 | 0·93 (0·86 – 1·01); p=0·068 | 0·90 (0·84 – 0·96); p=0·002 | 0·91 (0·84 – 0·97); p=0·0073 | 0·94 (0·89 – 1·00); p=0·038 |
| Male sex assigned at birth | 0·71 (0·28 – 1·77); p=0·44 | 0·59 (0·26 – 1·38); p=0·22 | 0·90 (0·35 – 2·30); p=0·83 | 0·73 (0·29 – 1·87); p=0·5 | 1·76 (0·69 – 4·46); p=0·22 | 1·22 (0·60 – 2·49); p=0·57 |
| Height (cm) | 0·98 (0·93 – 1·03); p=0·41 | 0·99 (0·94 – 1·04); p=0·59 | 0·99 (0·94 – 1·04); p=0·69 | 0·98 (0·93 – 1·03); p=0·45 | 0·95 (0·90 – 1·00); p=0·052 | 0·98 (0·94 – 1·02); p=0·35 |
| Weight (kg) | 0·99 (0·97 – 1·01); p=0·34 | 1·00 (0·98 – 1·02); p=0·82 | 1·00 (0·98 – 1·02); p=0·99 | 0·99 (0·97 – 1·01); p=0·45 | 1·00 (0·98 – 1·02); p=0·7 | 1·00 (0·98 – 1·01); p=0·95 |
| Vaccine type used for primary vaccination (acellular Pertussis vs whole cell Pertussis [reference]) | 1·22 (0·48 – 3·10); p=0·66 | 1·73 (0·74 – 4·04); p=0·2 | 1·22 (0·48 – 3·09); p=0·67 | 2·42 (0·99 – 5·93); p=0·053 | 1·83 (0·72 – 4·68); p=0·2 | 1·53 (0·76 – 3·09); p=0·22 |
| Time since the last pertussis vaccine dose (years) | 0·98 (0·93 – 1·04); p=0·51 | 0·97 (0·92 – 1·02); p=0·25 | 0·95 (0·90 – 1·01); p=0·12 | 0·93 (0·89 – 0·98); p=0·0058 | 0·95 (0·90 – 1·00); p=0·045 | 0·98 (0·94 – 1·02); p=0·3 |
| **Fractional absolute avidity** |  |  |  |  |  |  |
| Clinical disease outcome (symptomatic vs asymptomatic [reference]) | 0·97 (0·34 – 2·75); p=0·95 | 1·19 (0·44 – 3·21); p=0·73 | 2·34 (0·76 – 7·21); p=0·13 | 2·23 (0·61 – 8·07); p=0·21 | 3·48 (1·15 – 10·55); p=0·029 | 2·16 (0·81 – 5·74); p=0·12 |
| Challenge dose (10⁷ *vs.* 5×10⁶ [reference] CFU) | 2·00 (0·33 – 11·98); p=0·43 | 1·65 (0·30 – 9·03); p=0·55 | 2·69 (0·36 – 20·01); p=0·32 | 1·32 (0·15 – 11·48); p=0·8 | 1·97 (0·25 – 15·53); p=0·5 | 1·52 (0·27 – 8·49); p=0·62 |
| Challenge dose (5×10⁷ *vs.* 5×10⁶ [reference] CFU) | 0·82 (0·09 – 7·34); p=0·85 | 0·58 (0·07 – 4·64); p=0·59 | 2·29 (0·17 – 30·23); p=0·51 | 0·72 (0·05 – 11·34); p=0·81 | 0·51 (0·04 – 6·07); p=0·58 | 0·42 (0·05 – 3·33); p=0·4 |
| Age (years) | 0·95 (0·84 – 1·06); p=0·32 | 0·92 (0·83 – 1·02); p=0·11 | 0·87 (0·77 – 0·99); p=0·039 | 0·80 (0·72 – 0·90); p=<0·001 | 0·82 (0·73 – 0·92); p=0·0018 | 0·87 (0·79 – 0·97); p=0·013 |
| Male sex assigned at birth | 0·49 (0·12 – 1·93); p=0·3 | 0·44 (0·12 – 1·61); p=0·21 | 1·28 (0·26 – 6·27); p=0·75 | 0·71 (0·13 – 3·84); p=0·68 | 1·85 (0·35 – 9·75); p=0·45 | 1·36 (0·34 – 5·46); p=0·65 |
| Height (cm) | 0·96 (0·89 – 1·05); p=0·36 | 0·97 (0·90 – 1·05); p=0·45 | 0·96 (0·87 – 1·05); p=0·31 | 0·96 (0·87 – 1·06); p=0·42 | 0·93 (0·84 – 1·02); p=0·13 | 0·96 (0·89 – 1·04); p=0·27 |
| Weight (kg) | 0·99 (0·96 – 1·02); p=0·66 | 1·00 (0·97 – 1·03); p=0·94 | 1·01 (0·97 – 1·04); p=0·72 | 1·00 (0·97 – 1·04); p=0·93 | 1·00 (0·97 – 1·04); p=0·92 | 1·01 (0·98 – 1·04); p=0·58 |
| Vaccine type used for primary vaccination (acellular Pertussis vs whole cell Pertussis [reference]) | 1·58 (0·39 – 6·36); p=0·51 | 2·33 (0·63 – 8·58); p=0·2 | 2·26 (0·48 – 10·70); p=0·29 | 7·23 (1·55 – 33·84); p=0·014 | 4·34 (0·89 – 21·24); p=0·069 | 2·71 (0·71 – 10·41); p=0·14 |
| Time since the last pertussis vaccine dose (years) | 0·94 (0·87 – 1·02); p=0·15 | 0·94 (0·87 – 1·01); p=0·084 | 0·90 (0·81 – 0·99); p=0·035 | 0·87 (0·80 – 0·93); p=<0·001 | 0·88 (0·81 – 0·97); p=0·0089 | 0·92 (0·85 – 1·00); p=0·044 |

For each comparison, the fold‑change, lower and upper confidence interval bounds for the fold‑change, and the p‑value are reported.

**Table S8.** Univariate analyses to determine association of clinical, vaccinal and demographic factors with immunological properties of high avidity fractions before challenge with *Bordetella pertussis* and different time point post challenge.

|  | **Visit 1 (Day -1)** | **Visit 2 (Day 14)** | **Visit 3 (Day 28)** | **Visit 4 (Day 56)** | **Visit 5 (Day 180)** | **Visit 6 (Day 365)** |
| --- | --- | --- | --- | --- | --- | --- |
|  | **Model estimate (95% CI); p-value** | **Model estimate (95% CI); p-value** | **Model estimate (95% CI); p-value** | **Model estimate (95% CI); p-value** | **Model estimate (95% CI); p-value** | **Model estimate (95% CI); p-value** |
| **Anti-PT IgG** |  |  |  |  |  |  |
| Clinical disease outcome (symptomatic vs asymptomatic [reference]) | 0·98 (0·86 – 1·10); p=0·69 | 0·98 (0·84 – 1·14); p=0·79 | 1·90 (1·06 – 3·39); p=0·032 | 1·99 (1·02 – 3·91); p=0·045 | 1·87 (1·11 – 3·14); p=0·02 | 1·78 (1·12 – 2·83); p=0·017 |
| Challenge dose (10⁷ *vs.* 5×10⁶ [reference] CFU) | 1·10 (0·89 – 1·36); p=0·37 | 1·05 (0·81 – 1·37); p=0·7 | 1·26 (0·42 – 3·79); p=0·67 | 1·17 (0·35 – 3·85); p=0·79 | 1·63 (0·61 – 4·37); p=0·32 | 1·29 (0·53 – 3·16); p=0·56 |
| Challenge dose (5×10⁷ *vs.* 5×10⁶ [reference] CFU) | 1·00 (0·77 – 1·30); p=1 | 1·22 (0·89 – 1·68); p=0·21 | 1·23 (0·30 – 5·04); p=0·77 | 1·25 (0·27 – 5·71); p=0·77 | 1·07 (0·33 – 3·47); p=0·91 | 0·90 (0·31 – 2·65); p=0·84 |
| Age (years) | 0·99 (0·98 – 1·00); p=0·12 | 0·99 (0·97 – 1·01); p=0·26 | 0·93 (0·87 – 1·00); p=0·04 | 0·91 (0·85 – 0·97); p=0·0041 | 0·93 (0·88 – 0·99); p=0·018 | 0·95 (0·90 – 1·00); p=0·047 |
| Male sex assigned at birth | 0·90 (0·77 – 1·06); p=0·18 | 1·03 (0·84 – 1·27); p=0·76 | 1·27 (0·55 – 2·98); p=0·56 | 0·85 (0·34 – 2·16); p=0·73 | 0·92 (0·42 – 2·04); p=0·84 | 1·05 (0·52 – 2·12); p=0·90 |
| Height (cm) | 1·00 (0·99 – 1·01); p=0·84 | 0·99 (0·98 – 1·01); p=0·29 | 0·98 (0·93 – 1·03); p=0·34 | 1·00 (0·95 – 1·06); p=0·85 | 1·00 (0·95 – 1·05); p=0·95 | 0·99 (0·95 – 1·03); p=0·75 |
| Weight (kg) | 1·00 (1·00 – 1·00); p=0·85 | 1·00 (0·99 – 1·00); p=0·31 | 1·01 (0·99 – 1·03); p=0·40 | 1·01 (0·99 – 1·03); p=0·21 | 1·01 (0·99 – 1·03); p=0·27 | 1·01 (1·00 – 1·02); p=0·14 |
| Vaccine type used for primary vaccination (acellular Pertussis vs whole cell Pertussis [reference]) | 1·14 (0·97 – 1·34); p=0·1 | 1·18 (0·96 – 1·43); p=0·10 | 2·42 (1·11 – 5·28); p=0·028 | 3·05 (1·31 – 7·06); p=0·011 | 2·28 (1·10 – 4·73); p=0·028 | 1·73 (0·88 – 3·40); p=0·11 |
| Time since the last pertussis vaccine dose (years) | 1·00 (0·99 – 1·01); p=0·44 | 1·00 (0·99 – 1·01); p=0·73 | 0·95 (0·90 – 1·01); p=0·095 | 0·95 (0·90 – 1·00); p=0·033 | 0·97 (0·92 – 1·01); p=0·16 | 0·98 (0·94 – 1·02); p=0·26 |
| **Fractional relative avidity** |  |  |  |  |  |  |
| Clinical disease outcome (symptomatic vs asymptomatic [reference]) | 0·95 (0·67 – 1·35); p=0·77 | 0·93 (0·68 – 1·27); p=0·63 | 1·80 (0·89 – 3·63); p=0·1 | 1·97 (0·93 – 4·16); p=0·074 | 1·74 (0·88 – 3·45); p=0·11 | 2·18 (1·11 – 4·27); p=0·025 |
| Challenge dose (10⁷ *vs.* 5×10⁶ [reference] CFU) | 1·30 (0·72 – 2·36); p=0·37 | 1·13 (0·66 – 1·92); p=0·65 | 1·27 (0·35 – 4·61); p=0·71 | 0·93 (0·25 – 3·41); p=0·91 | 2·59 (0·78 – 8·55); p=0·11 | 2·25 (0·66 – 7·71); p=0·19 |
| Challenge dose (5×10⁷ *vs.* 5×10⁶ [reference] CFU) | 1·00 (0·48 – 2·07); p=1·00 | 1·44 (0·75 – 2·76); p=0·27 | 1·18 (0·23 – 6·18); p=0·84 | 1·23 (0·23 – 6·49); p=0·8 | 1·59 (0·38 – 6·63); p=0·51 | 0·97 (0·22 – 4·28); p=0·96 |
| Age (years) | 0·97 (0·94 – 1·01); p=0·12 | 0·98 (0·95 – 1·01); p=0·25 | 0·92 (0·85 – 1·00); p=0·042 | 0·89 (0·83 – 0·96); p=0·0028 | 0·91 (0·85 – 0·98); p=0·017 | 0·91 (0·85 – 0·99); p=0·025 |
| Male sex assigned at birth | 0·74 (0·47 – 1·17); p=0·19 | 1·02 (0·67 – 1·55); p=0·92 | 1·27 (0·47 – 3·44); p=0·63 | 0·64 (0·23 – 1·74); p=0·37 | 0·64 (0·24 – 1·69); p=0·35 | 0·81 (0·30 – 2·22); p=0·67 |
| Height (cm) | 1·00 (0·98 – 1·03); p=0·86 | 0·99 (0·97 – 1·01); p=0·39 | 0·97 (0·92 – 1·03); p=0·32 | 1·02 (0·96 – 1·08); p=0·57 | 1·00 (0·95 – 1·07); p=0·86 | 1·00 (0·94 – 1·06); p=0·99 |
| Weight (kg) | 1·00 (0·99 – 1·01); p=0·86 | 1·00 (0·99 – 1·01); p=0·39 | 1·00 (0·98 – 1·03); p=0·67 | 1·01 (0·99 – 1·03); p=0·55 | 1·01 (0·99 – 1·03); p=0·42 | 1·01 (0·99 – 1·03); p=0·37 |
| Vaccine type used for primary vaccination (acellular Pertussis vs whole cell Pertussis [reference]) | 1·44 (0·92 – 2·25); p=0·10 | 1·39 (0·93 – 2·08); p=0·1 | 2·77 (1·11 – 6·94); p=0·031 | 3·33 (1·33 – 8·37); p=0·012 | 2·61 (1·04 – 6·56); p=0·042 | 2·90 (1·15 – 7·29); p=0·026 |
| Time since the last pertussis vaccine dose (years) | 0·99 (0·96 – 1·02); p=0·41 | 1·00 (0·97 – 1·02); p=0·76 | 0·95 (0·89 – 1·02); p=0·15 | 0·94 (0·89 – 0·99); p=0·03 | 0·96 (0·91 – 1·02); p=0·17 | 0·97 (0·91 – 1·03); p=0·25 |
| **Fractional absolute avidity** |  |  |  |  |  |  |
| Clinical disease outcome (symptomatic vs asymptomatic [reference]) | 1·11 (0·57 – 2·17); p=0·75 | 1·00 (0·50 – 1·99); p=0·99 | 3·37 (1·01 – 11·24); p=0·048 | 3·53 (0·90 – 13·87); p=0·069 | 3·89 (1·23 – 12·36); p=0·023 | 4·12 (1·32 – 12·83); p=0·017 |
| Challenge dose (10⁷ *vs.* 5×10⁶ [reference] CFU) | 2·23 (0·75 – 6·64); p=0·14 | 1·46 (0·44 – 4·84); p=0·53 | 2·30 (0·25 – 21·47); p=0·45 | 1·39 (0·13 – 15·17); p=0·78 | 5·09 (0·63 – 40·98); p=0·12 | 3·67 (0·46 – 29·28); p=0·21 |
| Challenge dose (5×10⁷ *vs.* 5×10⁶ [reference] CFU) | 0·87 (0·23 – 3·29); p=0·83 | 1·08 (0·25 – 4·70); p=0·92 | 1·87 (0·11 – 32·84); p=0·66 | 1·41 (0·07 – 29·81); p=0·82 | 1·03 (0·08 – 12·40); p=0·98 | 0·61 (0·05 – 7·49); p=0·69 |
| Age (years) | 0·94 (0·88 – 1·01); p=0·10 | 0·95 (0·88 – 1·02); p=0·17 | 0·86 (0·75 – 0·99); p=0·041 | 0·80 (0·71 – 0·91); p=0·0011 | 0·83 (0·73 – 0·94); p=0·0047 | 0·85 (0·75 – 0·97); p=0·014 |
| Male sex assigned at birth | 0·52 (0·22 – 1·22); p=0·13 | 0·76 (0·30 – 1·89); p=0·54 | 1·80 (0·32 – 10·23); p=0·49 | 0·62 (0·10 – 3·95); p=0·60 | 0·67 (0·12 – 3·91); p=0·65 | 0·90 (0·16 – 5·07); p=0·9 |
| Height (cm) | 0·99 (0·94 – 1·04); p=0·64 | 0·97 (0·92 – 1·03); p=0·33 | 0·94 (0·85 – 1·04); p=0·2 | 1·00 (0·90 – 1·11); p=0·97 | 0·98 (0·89 – 1·09); p=0·75 | 0·98 (0·89 – 1·08); p=0·62 |
| Weight (kg) | 1·00 (0·98 – 1·02); p=0·7 | 1·00 (0·98 – 1·02); p=0·94 | 1·01 (0·97 – 1·05); p=0·57 | 1·02 (0·98 – 1·06); p=0·43 | 1·01 (0·98 – 1·05); p=0·45 | 1·02 (0·98 – 1·05); p=0·31 |
| Vaccine type used for primary vaccination (acellular Pertussis vs whole cell Pertussis [reference]) | 1·86 (0·78 – 4·43); p=0·16 | 1·88 (0·76 – 4·60); p=0·16 | 5·15 (1·01 – 26·23); p=0·049 | 9·95 (1·87 – 52·85); p=0·0089 | 6·18 (1·22 – 31·22); p=0·029 | 5·13 (1·02 – 25·68); p=0·047 |
| Time since the last pertussis vaccine dose (years) | 0·95 (0·90 – 1·00); p=0·044 | 0·96 (0·91 – 1·01); p=0·12 | 0·90 (0·80 – 1·01); p=0·062 | 0·87 (0·80 – 0·96); p=0·0051 | 0·90 (0·81 – 0·99); p=0·033 | 0·91 (0·82 – 1·01); p=0·063 |

For each comparison, the fold‑change, lower and upper confidence interval bounds for the fold‑change, and the p‑value are reported.

**Table S9.** Univariate analyses to determine association of clinical, vaccinal and demographic factors with immunological properties of very high avidity fractions before challenge with *Bordetella pertussis* and different time point post challenge.

|  | **Visit 1 (Day -1)** | **Visit 2 (Day 14)** | **Visit 3 (Day 28)** | **Visit 4 (Day 56)** | **Visit 5 (Day 180)** | **Visit 6 (Day 365)** |
| --- | --- | --- | --- | --- | --- | --- |
|  | **Model estimate (95% CI); p-value** | **Model estimate (95% CI); p-value** | **Model estimate (95% CI); p-value** | **Model estimate (95% CI); p-value** | **Model estimate (95% CI); p-value** | **Model estimate (95% CI); p-value** |
| **Anti-PT IgG** |  |  |  |  |  |  |
| Clinical disease outcome (symptomatic vs asymptomatic [reference]) | 1·00 (1·00 – 1·00); p=0·49 | 1·00 (1·00 – 1·00); p=0·49 | 1·25 (0·96 – 1·63); p=0·091 | 1·32 (0·94 – 1·86); p=0·10 | 1·21 (0·94 – 1·57); p=0·13 | 1·05 (0·92 – 1·20); p=0·47 |
| Challenge dose (10⁷ *vs.* 5×10⁶ [reference] CFU) | 1·00 (1·00 – 1·00); p=0·054 | 1·00 (1·00 – 1·00); p=0·054 | 1·00 (0·61 – 1·62); p=0·99 | 1·01 (0·56 – 1·82); p=0·98 | 0·84 (0·53 – 1·34); p=0·45 | 0·82 (0·66 – 1·02); p=0·068 |
| Challenge dose (5×10⁷ *vs.* 5×10⁶ [reference] CFU) | 1·00 (1·00 – 1·00); p=0·11 | 1·00 (1·00 – 1·00); p=0·11 | 0·95 (0·51 – 1·77); p=0·87 | 0·91 (0·43 – 1·92); p=0·79 | 0·84 (0·48 – 1·47); p=0·54 | 0·82 (0·63 – 1·06); p=0·13 |
| Age (years) | 1·00 (1·00 – 1·00); p=0·82 | 1·00 (1·00 – 1·00); p=0·82 | 0·97 (0·94 – 1·00); p=0·081 | 0·97 (0·94 – 1·00); p=0·085 | 0·98 (0·95 – 1·01); p=0·22 | 1·00 (0·98 – 1·01); p=0·81 |
| Male sex assigned at birth | 1·00 (1·00 – 1·00); p=0·29 | 1·00 (1·00 – 1·00); p=0·29 | 1·05 (0·72 – 1·52); p=0·81 | 1·08 (0·69 – 1·72); p=0·72 | 1·31 (0·92 – 1·87); p=0·12 | 1·10 (0·92 – 1·31); p=0·29 |
| Height (cm) | 1·00 (1·00 – 1·00); p=0·62 | 1·00 (1·00 – 1·00); p=0·62 | 1·00 (0·98 – 1·02); p=0·79 | 1·00 (0·98 – 1·03); p=0·89 | 1·00 (0·98 – 1·02); p=0·76 | 1·00 (0·99 – 1·01); p=0·64 |
| Weight (kg) | 1·00 (1·00 – 1·00); p=<0·001 | 1·00 (1·00 – 1·00); p=<0·001 | 1·01 (1·00 – 1·01); p=0·15 | 1·01 (1·00 – 1·02); p=0·21 | 1·01 (1·00 – 1·01); p=0·034 | 1·01 (1·00 – 1·01); p=<0·001 |
| Vaccine type used for primary vaccination (acellular Pertussis vs whole cell Pertussis [reference]) | 1·00 (1·00 – 1·00); p=0·39 | 1·00 (1·00 – 1·00); p=0·39 | 1·33 (0·93 – 1·90); p=0·12 | 1·57 (1·02 – 2·43); p=0·041 | 1·15 (0·80 – 1·66); p=0·44 | 0·93 (0·77 – 1·11); p=0·40 |
| Time since the last pertussis vaccine dose (years) | 1·00 (1·00 – 1·00); p=0·66 | 1·00 (1·00 – 1·00); p=0·66 | 0·98 (0·96 – 1·00); p=0·093 | 0·98 (0·95 – 1·00); p=0·10 | 0·99 (0·96 – 1·01); p=0·22 | 1·00 (0·99 – 1·01); p=0·66 |
| **Fractional relative avidity** |  |  |  |  |  |  |
| Clinical disease outcome (symptomatic vs asymptomatic [reference]) | 1·00 (1·00 – 1·00); p=0·49 | 1·00 (1·00 – 1·00); p=0·49 | 1·30 (0·96 – 1·75); p=0·087 | 1·32 (0·94 – 1·86); p=0·1 | 1·28 (0·94 – 1·75); p=0·11 | 1·05 (0·91 – 1·22); p=0·47 |
| Challenge dose (10⁷ *vs.* 5×10⁶ [reference] CFU) | 1·00 (1·00 – 1·00); p=0·054 | 1·00 (1·00 – 1·00); p=0·054 | 1·01 (0·58 – 1·75); p=0·97 | 1·16 (0·65 – 2·08); p=0·6 | 0·90 (0·51 – 1·60); p=0·7 | 0·80 (0·63 – 1·02); p=0·068 |
| Challenge dose (5×10⁷ *vs.* 5×10⁶ [reference] CFU) | 1·00 (1·00 – 1·00); p=0·11 | 1·00 (1·00 – 1·00); p=0·11 | 0·95 (0·47 – 1·92); p=0·88 | 1·02 (0·49 – 2·14); p=0·95 | 0·88 (0·44 – 1·75); p=0·71 | 0·80 (0·59 – 1·07); p=0·13 |
| Age (years) | 1·00 (1·00 – 1·00); p=0·82 | 1·00 (1·00 – 1·00); p=0·82 | 0·97 (0·94 – 1·00); p=0·076 | 0·97 (0·93 – 1·00); p=0·065 | 0·97 (0·94 – 1·01); p=0·16 | 1·00 (0·98 – 1·02); p=0·81 |
| Male sex assigned at birth | 1·00 (1·00 – 1·00); p=0·29 | 1·00 (1·00 – 1·00); p=0·29 | 1·07 (0·70 – 1·64); p=0·74 | 0·99 (0·63 – 1·57); p=0·98 | 1·33 (0·86 – 2·06); p=0·19 | 1·11 (0·91 – 1·35); p=0·29 |
| Height (cm) | 1·00 (1·00 – 1·00); p=0·62 | 1·00 (1·00 – 1·00); p=0·62 | 1·00 (0·97 – 1·02); p=0·79 | 1·00 (0·97 – 1·03); p=0·96 | 0·99 (0·97 – 1·02); p=0·67 | 1·00 (0·99 – 1·01); p=0·64 |
| Weight (kg) | 1·00 (1·00 – 1·00); p=<0·001 | 1·00 (1·00 – 1·00); p=<0·001 | 1·01 (1·00 – 1·01); p=0·17 | 1·00 (0·99 – 1·01); p=0·62 | 1·01 (1·00 – 1·02); p=0·10 | 1·01 (1·00 – 1·01); p=<0·001 |
| Vaccine type used for primary vaccination (acellular Pertussis vs whole cell Pertussis [reference]) | 1·00 (1·00 – 1·00); p=0·39 | 1·00 (1·00 – 1·00); p=0·39 | 1·39 (0·93 – 2·09); p=0·11 | 1·74 (1·15 – 2·62); p=0·01 | 1·30 (0·83 – 2·03); p=0·24 | 0·92 (0·75 – 1·12); p=0·40 |
| Time since the last pertussis vaccine dose (years) | 1·00 (1·00 – 1·00); p=0·66 | 1·00 (1·00 – 1·00); p=0·66 | 0·98 (0·95 – 1·00); p=0·088 | 0·98 (0·95 – 1·00); p=0·091 | 0·98 (0·96 – 1·01); p=0·18 | 1·00 (0·98 – 1·01); p=0·66 |
| **Fractional absolute avidity** |  |  |  |  |  |  |
| Clinical disease outcome (symptomatic vs asymptomatic [reference]) | 1·17 (0·74 – 1·83); p=0·49 | 1·07 (0·66 – 1·74); p=0·77 | 2·43 (1·10 – 5·36); p=0·029 | 2·38 (0·92 – 6·13); p=0·072 | 2·87 (1·34 – 6·17); p=0·0087 | 1·99 (1·08 – 3·67); p=0·028 |
| Challenge dose (10⁷ *vs.* 5×10⁶ [reference] CFU) | 1·71 (0·82 – 3·58); p=0·15 | 1·29 (0·57 – 2·93); p=0·52 | 1·83 (0·41 – 8·09); p=0·41 | 1·74 (0·34 – 8·94); p=0·49 | 1·77 (0·41 – 7·62); p=0·43 | 1·30 (0·43 – 3·94); p=0·63 |
| Challenge dose (5×10⁷ *vs.* 5×10⁶ [reference] CFU) | 0·87 (0·35 – 2·13); p=0·74 | 0·75 (0·28 – 2·05); p=0·56 | 1·50 (0·22 – 10·13); p=0·67 | 1·17 (0·15 – 9·47); p=0·88 | 0·57 (0·10 – 3·27); p=0·51 | 0·51 (0·13 – 1·92); p=0·30 |
| Age (years) | 0·97 (0·92 – 1·02); p=0·23 | 0·97 (0·92 – 1·02); p=0·22 | 0·91 (0·83 – 1·00); p=0·049 | 0·87 (0·79 – 0·95); p=0·0029 | 0·88 (0·81 – 0·96); p=0·0078 | 0·93 (0·86 – 0·99); p=0·03 |
| Male sex assigned at birth | 0·70 (0·38 – 1·26); p=0·22 | 0·74 (0·39 – 1·39); p=0·34 | 1·52 (0·48 – 4·85); p=0·46 | 0·96 (0·26 – 3·49); p=0·95 | 1·40 (0·42 – 4·66); p=0·57 | 1·24 (0·50 – 3·06); p=0·63 |
| Height (cm) | 0·99 (0·95 – 1·02); p=0·41 | 0·98 (0·95 – 1·02); p=0·41 | 0·96 (0·90 – 1·03); p=0·24 | 0·98 (0·91 – 1·06); p=0·63 | 0·97 (0·91 – 1·05); p=0·44 | 0·98 (0·93 – 1·03); p=0·41 |
| Weight (kg) | 1·00 (0·99 – 1·02); p=0·66 | 1·00 (0·99 – 1·02); p=0·65 | 1·01 (0·99 – 1·04); p=0·33 | 1·01 (0·98 – 1·04); p=0·40 | 1·01 (0·99 – 1·04); p=0·29 | 1·02 (1·00 – 1·03); p=0·094 |
| Vaccine type used for primary vaccination (acellular Pertussis vs whole cell Pertussis [reference]) | 1·29 (0·70 – 2·36); p=0·40 | 1·35 (0·71 – 2·55); p=0·35 | 2·59 (0·85 – 7·83); p=0·09 | 5·19 (1·66 – 16·26); p=0·0064 | 3·08 (0·99 – 9·53); p=0·051 | 1·63 (0·66 – 3·98); p=0·27 |
| Time since the last pertussis vaccine dose (years) | 0·96 (0·93 – 0·99); p=0·018 | 0·96 (0·93 – 1·00); p=0·041 | 0·92 (0·86 – 0·99); p=0·027 | 0·91 (0·85 – 0·97); p=0·0036 | 0·92 (0·86 – 0·98); p=0·012 | 0·94 (0·89 – 0·99); p=0·018 |

For each comparison, the fold‑change, lower and upper confidence interval bounds for the fold‑change, and the p‑value are reported.

**Table S10.** Multivariate analyses to determine association of clinical disease outcome and age with immunological properties of low avidity fractions before challenge with *Bordetella pertussis* and different time point post challenge.

|  | **Visit 1 (Day -1)** | **Visit 2 (Day 14)** | **Visit 3 (Day 28)** | **Visit 4 (Day 56)** | **Visit 5 (Day 180)** | **Visit 6 (Day 365)** |
| --- | --- | --- | --- | --- | --- | --- |
|  | **Model estimate (95% CI); p-value** | **Model estimate (95% CI); p-value** | **Model estimate (95% CI); p-value** | **Model estimate (95% CI); p-value** | **Model estimate (95% CI); p-value** | **Model estimate (95% CI); p-value** |
| **Anti-PT IgG** |  |  |  |  |  |  |
| Clinical disease outcome (symptomatic vs asymptomatic [reference]) | 1·10 (0·49 – 2·50); p=0·8 | 1·01 (0·65 – 1·55); p=0·98 | 1·05 (0·63 – 1·76); p=0·84 | 1·25 (0·69 – 2·25); p=0·44 | 1·38 (0·84 – 2·27); p=0·19 | 1·53 (0·94 – 2·48); p=0·084 |
| Age (years) | 1·03 (0·93 – 1·14); p=0·49 | 0·97 (0·93 – 1·02); p=0·23 | 1·02 (0·97 – 1·08); p=0·4 | 0·96 (0·90 – 1·02); p=0·15 | 0·96 (0·91 – 1·02); p=0·22 | 1·00 (0·94 – 1·06); p=0·99 |
| **Fractional relative avidity** | |  |  |  |  |  |
| Clinical disease outcome (symptomatic vs asymptomatic [reference]) | 1·01 (0·47 – 2·18); p=0·97 | 0·95 (0·40 – 2·25); p=0·9 | 0·94 (0·46 – 1·93); p=0·86 | 0·77 (0·40 – 1·49); p=0·43 | 0·92 (0·49 – 1·74); p=0·79 | 1·22 (0·66 – 2·28); p=0·51 |
| Age (years) | 1·01 (0·93 – 1·10); p=0·86 | 0·96 (0·87 – 1·06); p=0·39 | 1·08 (0·99 – 1·18); p=0·064 | 0·97 (0·91 – 1·04); p=0·42 | 1·00 (0·93 – 1·08); p=0·97 | 0·98 (0·91 – 1·05); p=0·54 |
| **Fractional absolute avidity** | |  |  |  |  |  |
| Clinical disease outcome (symptomatic vs asymptomatic [reference]) | 1·14 (0·38 – 3·43); p=0·81 | 0·98 (0·28 – 3·39); p=0·97 | 1·61 (0·67 – 3·90); p=0·27 | 1·07 (0·48 – 2·38); p=0·86 | 1·86 (0·83 – 4·19); p=0·13 | 2·11 (0·79 – 5·65); p=0·13 |
| Age (years) | 0·98 (0·87 – 1·11); p=0·74 | 0·93 (0·81 – 1·07); p=0·29 | 1·03 (0·93 – 1·14); p=0·57 | 0·88 (0·81 – 0·96); p=0·0041 | 0·92 (0·84 – 1·01); p=0·072 | 0·92 (0·82 – 1·03); p=0·12 |

For each comparison, the fold‑change, lower and upper bounds of the confidence interval, and the p‑value are reported.

**Table S11.** Multivariate analyses to determine association of clinical disease outcome and age with immunological properties of low-medium avidity fractions before challenge with *Bordetella pertussis* and different time point post challenge.

|  | **Visit 1 (Day -1)** | **Visit 2 (Day 14)** | **Visit 3 (Day 28)** | **Visit 4 (Day 56)** | **Visit 5 (Day 180)** | **Visit 6 (Day 365)** |
| --- | --- | --- | --- | --- | --- | --- |
|  | **Model estimate (95% CI); p-value** | **Model estimate (95% CI); p-value** | **Model estimate (95% CI); p-value** | **Model estimate (95% CI); p-value** | **Model estimate (95% CI); p-value** | **Model estimate (95% CI); p-value** |
| **Anti-PT IgG** |  |  |  |  |  |  |
| Clinical disease outcome (symptomatic vs asymptomatic [reference]) | 1·04 (0·67 – 1·62); p=0·85 | 0·94 (0·48 – 1·87); p=0·86 | 1·71 (0·90 – 3·24); p=0·097 | 1·43 (0·75 – 2·72); p=0·27 | 2·01 (1·21 – 3·34); p=0·0092 | 1·66 (1·02 – 2·71); p=0·041 |
| Age (years) | 0·98 (0·94 – 1·03); p=0·49 | 1·04 (0·95 – 1·14); p=0·36 | 0·92 (0·86 – 0·99); p=0·035 | 0·90 (0·84 – 0·96); p=0·0041 | 0·91 (0·86 – 0·97); p=0·0041 | 0·93 (0·88 – 0·98); p=0·012 |
| **Fractional relative avidity** |  |  |  |  |  |  |
| Clinical disease outcome (symptomatic vs asymptomatic [reference]) | 1·21 (0·65 – 2·24); p=0·53 | 1·02 (0·62 – 1·66); p=0·94 | 1·10 (0·56 – 2·14); p=0·78 | 0·71 (0·38 – 1·36); p=0·29 | 1·40 (0·77 – 2·54); p=0·26 | 1·13 (0·67 – 1·92); p=0·63 |
| Age (years) | 1·01 (0·94 – 1·08); p=0·87 | 0·96 (0·91 – 1·02); p=0·18 | 0·94 (0·87 – 1·02); p=0·12 | 0·99 (0·92 – 1·05); p=0·66 | 0·97 (0·91 – 1·04); p=0·39 | 0·95 (0·90 – 1·01); p=0·1 |
| **Fractional absolute avidity** |  |  |  |  |  |  |
| Clinical disease outcome (symptomatic vs asymptomatic [reference]) | 1·36 (0·49 – 3·77); p=0·54 | 1·05 (0·43 – 2·57); p=0·92 | 1·88 (0·68 – 5·20); p=0·21 | 0·99 (0·37 – 2·66); p=0·99 | 2·83 (1·16 – 6·89); p=0·024 | 1·95 (0·87 – 4·39); p=0·1 |
| Age (years) | 0·98 (0·87 – 1·09); p=0·69 | 0·93 (0·85 – 1·03); p=0·17 | 0·90 (0·79 – 1·01); p=0·067 | 0·89 (0·80 – 0·99); p=0·031 | 0·89 (0·80 – 0·99); p=0·028 | 0·89 (0·81 – 0·98); p=0·016 |

For each comparison, the fold‑change, lower and upper bounds of the confidence interval, and the p‑value are reported.

**Table S12.** Multivariate analyses to determine association of clinical disease outcome and age with immunological properties of medium avidity fractions before challenge with *Bordetella pertussis* and different time point post challenge.

|  | **Visit 1 (Day -1)** | **Visit 2 (Day 14)** | **Visit 3 (Day 28)** | **Visit 4 (Day 56)** | **Visit 5 (Day 180)** | **Visit 6 (Day 365)** |
| --- | --- | --- | --- | --- | --- | --- |
|  | **Model estimate (95% CI); p-value** | **Model estimate (95% CI); p-value** | **Model estimate (95% CI); p-value** | **Model estimate (95% CI); p-value** | **Model estimate (95% CI); p-value** | **Model estimate (95% CI); p-value** |
| **Anti-PT IgG** |  |  |  |  |  |  |
| Clinical disease outcome (symptomatic vs asymptomatic [reference]) | 0·98 (0·67 – 1·42); p=0·89 | 1·02 (0·70 – 1·48); p=0·92 | 1·68 (0·89 – 3·17); p=0·11 | 1·57 (0·80 – 3·10); p=0·18 | 1·83 (1·06 – 3·16); p=0·031 | 1·61 (0·97 – 2·66); p=0·066 |
| Age (years) | 0·97 (0·94 – 1·02); p=0·21 | 0·97 (0·94 – 1·02); p=0·22 | 0·93 (0·86 – 1·00); p=0·053 | 0·89 (0·83 – 0·96); p=0·0027 | 0·92 (0·86 – 0·98); p=0·0089 | 0·94 (0·88 – 0·99); p=0·026 |
| **Fractional relative avidity** |  |  |  |  |  |  |
| Clinical disease outcome (symptomatic vs asymptomatic [reference]) | 1·19 (0·59 – 2·40); p=0·62 | 1·14 (0·56 – 2·35); p=0·7 | 1·19 (0·74 – 1·89); p=0·46 | 1·27 (0·75 – 2·17); p=0·36 | 1·07 (0·59 – 1·95); p=0·82 | 1·06 (0·54 – 2·07); p=0·86 |
| Age (years) | 0·96 (0·89 – 1·04); p=0·29 | 0·97 (0·90 – 1·05); p=0·48 | 0·96 (0·91 – 1·01); p=0·14 | 0·95 (0·89 – 1·00); p=0·05 | 0·96 (0·90 – 1·03); p=0·23 | 0·94 (0·87 – 1·02); p=0·11 |
| **Fractional absolute avidity** |  |  |  |  |  |  |
| Clinical disease outcome (symptomatic vs asymptomatic [reference]) | 1·34 (0·46 – 3·85); p=0·58 | 1·18 (0·38 – 3·65); p=0·77 | 2·03 (0·81 – 5·12); p=0·13 | 1·77 (0·65 – 4·86); p=0·25 | 2·16 (0·80 – 5·85); p=0·12 | 1·82 (0·65 – 5·13); p=0·24 |
| Age (years) | 0·93 (0·83 – 1·05); p=0·24 | 0·94 (0·83 – 1·07); p=0·34 | 0·91 (0·82 – 1·02); p=0·097 | 0·85 (0·77 – 0·95); p=0·0055 | 0·88 (0·78 – 0·99); p=0·03 | 0·88 (0·78 – 0·99); p=0·036 |

For each comparison, the fold‑change, lower and upper bounds of the confidence interval, and the p‑value are reported.

**Table S13.** Multivariate analyses to determine association of clinical disease outcome and age with immunological properties of medium-high avidity fractions before challenge with *Bordetella pertussis* and different time point post challenge.

|  | **Visit 1 (Day -1)** | **Visit 2 (Day 14)** | **Visit 3 (Day 28)** | **Visit 4 (Day 56)** | **Visit 5 (Day 180)** | **Visit 6 (Day 365)** |
| --- | --- | --- | --- | --- | --- | --- |
|  | **Model estimate (95% CI); p-value** | **Model estimate (95% CI); p-value** | **Model estimate (95% CI); p-value** | **Model estimate (95% CI); p-value** | **Model estimate (95% CI); p-value** | **Model estimate (95% CI); p-value** |
| **Anti-PT IgG** |  |  |  |  |  |  |
| Clinical disease outcome (symptomatic vs asymptomatic [reference]) | 0·91 (0·66 – 1·26); p=0·57 | 0·99 (0·75 – 1·32); p=0·96 | 1·59 (0·86 – 2·96); p=0·13 | 1·52 (0·78 – 2·96); p=0·21 | 1·88 (1·12 – 3·13); p=0·019 | 1·60 (0·95 – 2·71); p=0·075 |
| Age (years) | 0·98 (0·95 – 1·02); p=0·29 | 0·98 (0·94 – 1·01); p=0·11 | 0·93 (0·87 – 1·00); p=0·05 | 0·89 (0·83 – 0·96); p=0·0025 | 0·92 (0·86 – 0·97); p=0·0053 | 0·94 (0·89 – 1·00); p=0·051 |
| **Fractional relative avidity** |  |  |  |  |  |  |
| Clinical disease outcome (symptomatic vs asymptomatic [reference]) | 0·80 (0·39 – 1·61); p=0·51 | 1·03 (0·54 – 1·95); p=0·93 | 1·10 (0·56 – 2·17); p=0·77 | 0·94 (0·49 – 1·80); p=0·85 | 1·39 (0·76 – 2·55); p=0·27 | 1·05 (0·64 – 1·74); p=0·83 |
| Age (years) | 0·97 (0·90 – 1·05); p=0·43 | 0·95 (0·88 – 1·02); p=0·13 | 0·93 (0·86 – 1·01); p=0·089 | 0·90 (0·84 – 0·96); p=0·0029 | 0·91 (0·85 – 0·98); p=0·011 | 0·94 (0·89 – 1·00); p=0·047 |
| **Fractional absolute avidity** |  |  |  |  |  |  |
| Clinical disease outcome (symptomatic vs asymptomatic [reference]) | 0·90 (0·31 – 2·58); p=0·83 | 1·06 (0·40 – 2·83); p=0·91 | 1·89 (0·63 – 5·70); p=0·25 | 1·31 (0·44 – 3·89); p=0·61 | 2·82 (1·10 – 7·20); p=0·032 | 1·82 (0·73 – 4·50); p=0·19 |
| Age (years) | 0·94 (0·84 – 1·06); p=0·32 | 0·92 (0·82 – 1·02); p=0·12 | 0·89 (0·78 – 1·01); p=0·071 | 0·81 (0·72 – 0·91); p=<0·001 | 0·83 (0·75 – 0·93); p=0·0021 | 0·88 (0·80 – 0·98); p=0·021 |

For each comparison, the fold‑change, lower and upper bounds of the confidence interval, and the p‑value are reported.

**Table S14.** Multivariate analyses to determine association of clinical disease outcome and age with immunological properties of high avidity fractions before challenge with *Bordetella pertussis* and different time point post challenge.

|  | **Visit 1 (Day -1)** | **Visit 2 (Day 14)** | **Visit 3 (Day 28)** | **Visit 4 (Day 56)** | **Visit 5 (Day 180)** | **Visit 6 (Day 365)** |
| --- | --- | --- | --- | --- | --- | --- |
|  | **Model estimate (95% CI); p-value** | **Model estimate (95% CI); p-value** | **Model estimate (95% CI); p-value** | **Model estimate (95% CI); p-value** | **Model estimate (95% CI); p-value** | **Model estimate (95% CI); p-value** |
| **Anti-PT IgG** |  |  |  |  |  |  |
| Clinical disease outcome (symptomatic vs asymptomatic [reference]) | 0·96 (0·85 – 1·09); p=0·52 | 0·97 (0·83 – 1·13); p=0·66 | 1·71 (0·97 – 3·02); p=0·065 | 1·60 (0·85 – 3·00); p=0·14 | 1·73 (1·07 – 2·80); p=0·026 | 1·67 (1·06 – 2·62); p=0·027 |
| Age (years) | 0·99 (0·98 – 1·00); p=0·11 | 0·99 (0·97 – 1·01); p=0·25 | 0·94 (0·88 – 1·01); p=0·083 | 0·92 (0·86 – 0·98); p=0·012 | 0·94 (0·89 – 0·99); p=0·024 | 0·95 (0·91 – 1·01); p=0·076 |
| **Fractional relative avidity** |  |  |  |  |  |  |
| Clinical disease outcome (symptomatic vs asymptomatic [reference]) | 0·91 (0·65 – 1·29); p=0·59 | 0·90 (0·66 – 1·23); p=0·51 | 1·58 (0·79 – 3·15); p=0·19 | 1·52 (0·77 – 3·03); p=0·22 | 1·57 (0·84 – 2·97); p=0·15 | 1·96 (1·03 – 3·71); p=0·041 |
| Age (years) | 0·97 (0·93 – 1·01); p=0·11 | 0·98 (0·95 – 1·01); p=0·22 | 0·93 (0·86 – 1·01); p=0·079 | 0·90 (0·84 – 0·97); p=0·0081 | 0·92 (0·85 – 0·99); p=0·024 | 0·93 (0·86 – 1·00); p=0·041 |
| **Fractional absolute avidity** |  |  |  |  |  |  |
| Clinical disease outcome (symptomatic vs asymptomatic [reference]) | 1·03 (0·53 – 1·99); p=0·93 | 0·93 (0·47 – 1·85); p=0·83 | 2·71 (0·83 – 8·85); p=0·096 | 2·12 (0·63 – 7·15); p=0·21 | 3·19 (1·16 – 8·79); p=0·027 | 3·37 (1·17 – 9·71); p=0·026 |
| Age (years) | 0·94 (0·88 – 1·02); p=0·12 | 0·95 (0·88 – 1·02); p=0·17 | 0·89 (0·77 – 1·02); p=0·081 | 0·82 (0·72 – 0·93); p=0·0033 | 0·84 (0·75 – 0·95); p=0·0057 | 0·87 (0·77 – 0·98); p=0·023 |

For each comparison, the fold‑change, lower and upper bounds of the confidence interval, and the p‑value are reported.

**Table S15.** Multivariate analyses to determine association of clinical disease outcome and age with immunological properties of very high avidity fractions before challenge with *Bordetella pertussis* and different time point post challenge.

|  | **Visit 1 (Day -1)** | **Visit 2 (Day 14)** | **Visit 3 (Day 28)** | **Visit 4 (Day 56)** | **Visit 5 (Day 180)** | **Visit 6 (Day 365)** |
| --- | --- | --- | --- | --- | --- | --- |
|  | **Model estimate (95% CI); p-value** | **Model estimate (95% CI); p-value** | **Model estimate (95% CI); p-value** | **Model estimate (95% CI); p-value** | **Model estimate (95% CI); p-value** | **Model estimate (95% CI); p-value** |
| **Anti-PT IgG** |  |  |  |  |  |  |
| Clinical disease outcome (symptomatic vs asymptomatic [reference]) | 1·00 (1·00 – 1·00); p=0·51 | 1·00 (1·00 – 1·00); p=0·51 | 1·20 (0·92 – 1·57); p=0·16 | 1·24 (0·88 – 1·76); p=0·21 | 1·19 (0·92 – 1·54); p=0·17 | 1·05 (0·91 – 1·20); p=0·49 |
| Age (years) | 1·00 (1·00 – 1·00); p=0·9 | 1·00 (1·00 – 1·00); p=0·9 | 0·98 (0·95 – 1·01); p=0·14 | 0·98 (0·94 – 1·01); p=0·17 | 0·98 (0·96 – 1·01); p=0·29 | 1·00 (0·98 – 1·01); p=0·91 |
| **Fractional relative avidity** |  |  |  |  |  |  |
| Clinical disease outcome (symptomatic vs asymptomatic [reference]) | 1·00 (1·00 – 1·00); p=0·51 | 1·00 (1·00 – 1·00); p=0·51 | 1·24 (0·92 – 1·67); p=0·16 | 1·24 (0·88 – 1·74); p=0·21 | 1·25 (0·92 – 1·71); p=0·15 | 1·05 (0·90 – 1·23); p=0·49 |
| Age (years) | 1·00 (1·00 – 1·00); p=0·9 | 1·00 (1·00 – 1·00); p=0·9 | 0·97 (0·94 – 1·01); p=0·14 | 0·97 (0·94 – 1·01); p=0·14 | 0·98 (0·94 – 1·01); p=0·22 | 1·00 (0·98 – 1·02); p=0·91 |
| **Fractional absolute avidity** |  |  |  |  |  |  |
| Clinical disease outcome (symptomatic vs asymptomatic [reference]) | 1·13 (0·71 – 1·77); p=0·6 | 1·03 (0·63 – 1·67); p=0·91 | 2·12 (0·97 – 4·64); p=0·059 | 1·72 (0·72 – 4·11); p=0·21 | 2·53 (1·28 – 5·00); p=0·0095 | 1·82 (1·01 – 3·25); p=0·045 |
| Age (years) | 0·97 (0·93 – 1·02); p=0·27 | 0·97 (0·92 – 1·02); p=0·24 | 0·93 (0·85 – 1·02); p=0·099 | 0·88 (0·80 – 0·96); p=0·0082 | 0·90 (0·83 – 0·97); p=0·0086 | 0·94 (0·88 – 1·00); p=0·049 |

For each comparison, the fold‑change, lower and upper bounds of the confidence interval, and the p‑value are reported.

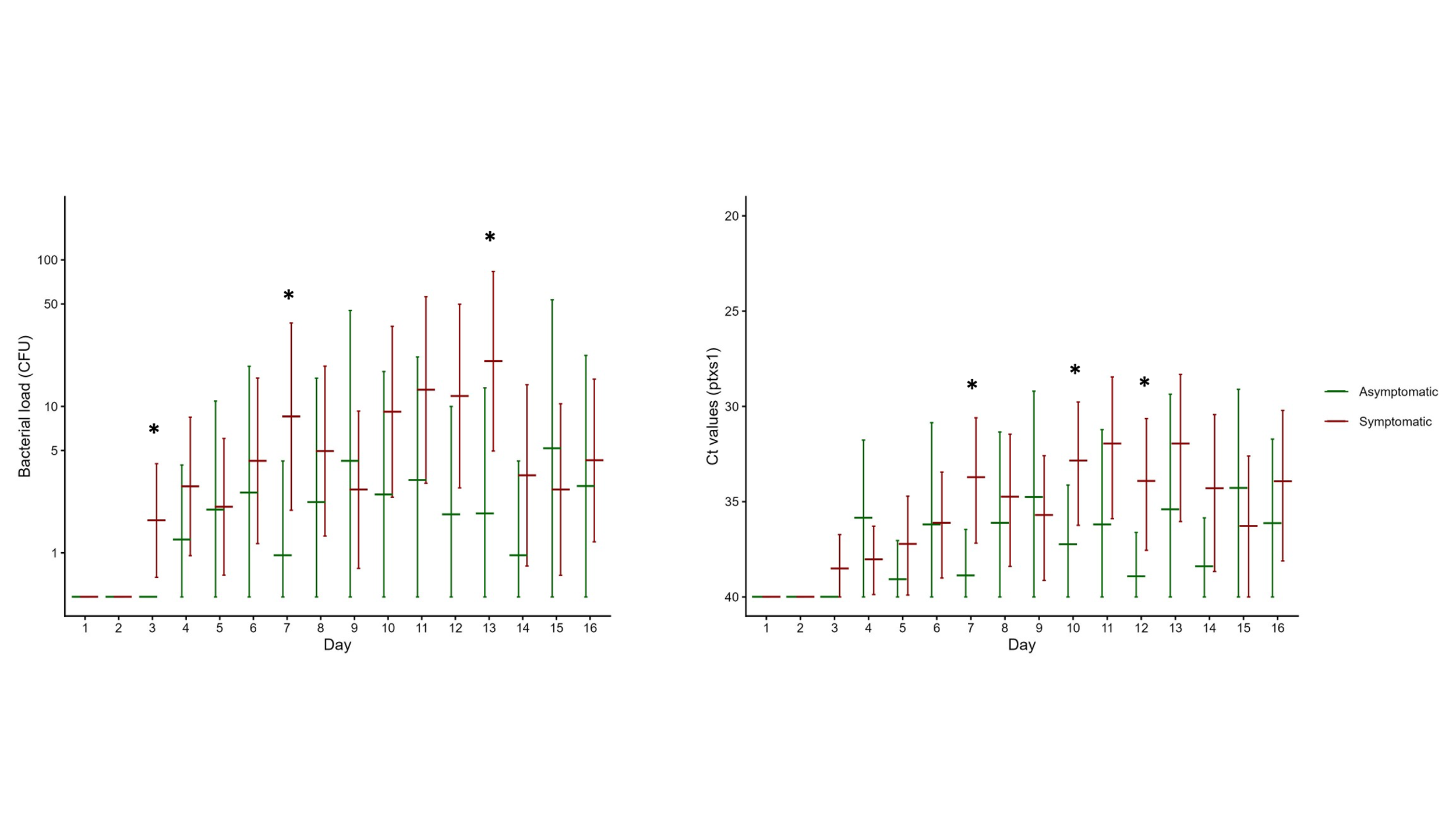

**Figure S1: Mucosal surface colonization following intranasal instillation with *Bordetella pertussis*. A)** Bacterial loads quantified by plating nasal samples collected —days 1 to 16— from asymptomatic (green) and symptomatic (red) participants infected with *B. pertussis*. **B)** Expression of the gene encoding pertussis toxin subunit 1 (*ptxs1*) measured by quantitative PCR. Samples with undetectable amplification were assigned a Ct value of 40 (the maximum number of cycles performed), corresponding to the threshold‑cycle definition (i.e., the number of amplification cycles required for the fluorescence signal to exceed assay threshold). Crosses represent the geometric mean (horizontal line) with its 95% confidence interval (error bar). Statistical comparisons were performed using unpaired Welch’s t test. *, *p* < 0.05.
